# Convergent molecular signatures across eating disorders and obsessive-compulsive disorder in the human brain

**DOI:** 10.1101/2024.11.27.24318078

**Authors:** Michael S. Breen, Ran Tao, Andy Yang, Xuran Wang, Pardis Amini, Miguel Rodriguez de los Santos, Anna C. Brandtjen, Amy Deep-Soboslay, Walter H. Kaye, Thomas M. Hyde, Joel E. Kleinman, Joseph D. Buxbaum, Dorothy E. Grice

## Abstract

Eating disorders (ED) and obsessive-compulsive disorder (OCD) exhibit significant clinical and genetic overlap, yet their shared molecular mechanisms remain unclear. We conducted a transcriptomic investigation of the dorsolateral prefrontal cortex (DLPFC) and caudate from 86 controls, 57 ED, and 27 OCD cases. ED was associated with robust differentially expressed genes (DEGs): 102 DEGs the DLPFC and 222 in the caudate (FDR < 1%) and replicated in an independent cohort. For OCD, no DEGs reached significance; however, meta-analysis with extant data identified 57 DEGs in the caudate. High concordance in transcriptomic changes was observed between ED and OCD in both regions (DLPFC *r*=0.67, caudate *r*=0.75). A combined ED+OCD analysis uncovered 233 DEGs in the DLPFC and 816 in the caudate, implicating disrupted GABAergic neuron function, neuroendocrine pathways, metabolism, and synaptic processes. Genetically regulated expression analysis identified nine genes with strong evidence for increasing ED risk, further validating these pathways. These findings reveal a shared molecular basis for ED and OCD, offering new insights into their pathobiology and potential therapeutic targets.

## INTRODUCTION

Eating disorders (ED) – particularly anorexia nervosa and bulimia nervosa - and obsessive-compulsive disorder (OCD) are complex neuropsychiatric conditions, each defined by distinct clinical manifestations yet sharing common transdiagnostic behaviors and motivations^1–6^. The genetic overlap between these disorders is increasingly supported by familial patterns^7,8^, twin studies^9,10^, and advanced genome-wide associations^11–17^, suggesting a shared compulsive genetic framework that contributes to overlapping obsessive-compulsive symptomatology. Moreover, epidemiological evidence points to a heightened comorbid risk^10,18^, hinting at a shared etiological framework. Although the evidence intertwining ED and OCD imply a common foundation, delineating their shared molecular pathways has been challenging.

Recent transcriptomic studies have begun to illuminate the operative genetic pathways involved in these conditions, uncovering notable dysregulation within crucial neurobiological circuits, particularly in the frontal cortex and striatum. In OCD, altered gene expression across striatal regions—specifically the caudate, nucleus accumbens, and putamen—points to perturbations in synaptic regulation, immune function, and neurotransmitter homeostasis, each region leaving a distinct transcriptomic signature^19–22^. In ED, preliminary findings have pointed to the dysregulation of genes within the dorsolateral prefrontal cortex (DLPFC), which are implicated in metabolic pathways also disturbed in OCD^22^, reinforcing the concept of a functional linkage between the disorders. Collectively, these insights, while promising, stem from studies that are limited by sample size, emphasizing the need for more comprehensive research to fully understand the shared biological foundations.

Clinically, ED and OCD are characterized by anxiety-provoking thoughts and related compulsive behaviors that serve to avoid certain stimuli or to reduce the feelings of distress associated with the difficult thoughts ^1,3,23^. While the themes of the thoughts and behaviors associated with ED and OCD are diagnostically distinct, these disorders share a common thread of anxiety-driven and ritualized response patterns^3^. Therapeutically, there is also overlap in the evidence-based treatments – the cognitive-behavioral therapy approaches and pharmacological treatments – for these disorders ^24–28^. This suggests underlying commonalities in the mechanisms of therapeutic action, despite the specific focus and intended effects of each treatment modality. The phenomenological and therapeutic overlap between ED and OCD also hints at shared molecular pathways and neural circuitry that have yet to be fully understood. A deeper insight into the shared and distinct molecular underpinnings in ED and OCD could support innovative treatment approaches and enable more effective interventions for individuals grappling with these difficult disorders.

This study, the largest of its kind to date, was designed to examine the transcriptomic profiles of the caudate and DLPFC in individuals with ED or OCD. These regions are integral to the cortico-striato-thalamic circuitry and executive function and posited as key anatomical regions in the manifestation of maladaptive behaviors seen in both disorders. To verify replication within the caudate, we employed an additional, smaller cohort. We also integrate our research with existing datasets and corroborate our results, deepen the understanding of the shared and distinct molecular pathways underlying ED and OCD and reveal new opportunities to support the development of novel therapeutics.

## METHODS

### Postmortem Tissue Acquisition and Clinical Characterization

Human brain tissue for this study was obtained from the Lieber Institute for Brain Development (LIBD) Human Brain and Tissue Repository, following rigorous ethical protocols across multiple acquisition sites. Ethics committee of LIBD gave ethical approval for this work. Tissue was collected at the time of autopsy from the Office of the Chief Medical Examiner of the State of Maryland, under Maryland Department of Health IRB protocol #12–24; the Departments of Pathology at Western Michigan University Homer Stryker MD School of Medicine and the University of North Dakota School of Medicine and Health Sciences; Gift of Life Michigan; and the County of Santa Clara Medical Examiner-Coroner Office, all under WCG IRB protocol #20111080. Additional samples were acquired through the National Institute of Mental Health Intramural Research Program (NIH protocol #90-M-0142) via material transfer agreement with LIBD.

Informed consent was obtained via audiotaped and witnessed documentation from the legal next-of-kin for all cases. A 39-item LIBD Autopsy telephone screening was conducted at the time of donation with the legal next-of-kin to gather data on the donor’s medical, psychiatric, social, and substance use history. Clinical diagnoses were retrospectively reviewed for every donor using information from autopsy reports, toxicology testing, forensic investigations, neuropathological examinations, telephone screenings, psychiatric/substance abuse treatment records, and supplemental family informant interviews. These data were compiled into detailed psychiatric narrative summaries, which were independently reviewed by two board-certified psychiatrists to establish lifetime psychiatric, and substance use diagnoses according to DSM-5 criteria.

Toxicology testing, conducted by medical examiners as part of autopsy and forensic investigations, included assessments for drugs of abuse (e.g., ethanol/volatiles, cocaine/metabolites, amphetamines, opiates). Additional postmortem toxicology testing was performed by National Medical Services (www.nmslabs.com), including assessments for nicotine/cotinine and cannabinoids for all cases, and for antidepressants, mood stabilizers, and antipsychotics in psychiatric cases, using blood and/or cerebellar tissue samples.

Non-psychiatric control donors had no lifetime history of psychiatric or substance use disorders, as determined by DSM-5 criteria, and were excluded if they presented with evidence of acute drug or alcohol intoxication at the time of death. All donors were excluded if neuropathological abnormalities such as cerebrovascular accidents, neuritic pathology, or other structural brain abnormalities were identified.

### RNA-sequencing data generation

#### Discovery cohort

We assembled a discovery RNA-seq cohort, comprising paired measurements of the DLPFC and caudate from 86 controls, 57 ED cases, and 27 OCD cases. The ED cases were comprised of eight anorexia nervosa restricting type (AN-R), 25 with binging and purging behaviors (AN-BP, *n*=14; bulimia nervosa, *n*=11), and the remaining were unspecified ED. Individuals in this cohort displayed varying biological sex (103 female, 66 male), age (41.5 ± 14.8 years) and ethnicity (**Supplemental Table 1**). Total RNA was extracted from all tissue samples using AllPrep DNA/RNA Mini Kit (Qiagen Cat No./ID: 80204) and displayed high RIN values (7.3 ± 1.3). Paired-end strand-specific sequencing libraries were prepared from 300ng total RNA using the TruSeq Stranded Total RNA Library Preparation kit with Ribo-Zero Gold ribosomal RNA depletion which removes rRNA and mtRNA. The libraries were then sequenced on an Illumina HiSeq 3000 at the Promega sequencing facility, producing a mean library depth of 17,299,335 reads across 200-bp paired-end reads per sample.

#### Replication cohort

To validate our findings, we generated a second, smaller RNA-seq replication cohort focusing on the caudate. Relative to the discovery cohort, this replication cohort comprised 41 ED cases, including 37 technical replicates and 4 unique biological replicates, 42 neurotypical controls with 30 technical replicates and 12 unique biological replicates, and 44 MDD non-ED neuropsychiatric controls. In this cohort, all individuals were female, of European descent, and of similar age (47.3 ±17.4 years) (**Supplemental Table 1**). RNA extraction and sequencing was conducted using matching protocols on high RIN samples (8.4 ± 0.9), but carried out at the LIBD Sequencing Facility, producing a mean library depth of 26,205,275 across 200-bp paired-end reads per sample.

### RNA-sequencing data processing and outlier removal

RNA-sequencing data were subjected to a rigorous processing protocol using our established pipeline (https://github.com/CommonMindConsortium/RAPiD-nf/). Initially, raw RNA sequences were aligned to the hg38 reference genome from Ensembl via the STAR algorithm^29^. Quality control checks, including assessments of GC content, duplication rates, gene body coverage, and library complexity were carried out using RSeQC^30^ and Picard tools (https://broadinstitute.github.io/picard/). These procedures also included unsupervised clustering and marking of duplicate reads. Gene expression levels were quantified with featureCounts^31^ and the resulting expression matrix was curated to retain genes present in at least one-third of our sample set. This matrix was then normalized through the VOOM method^32^.

#### Discovery cohort

A final normalized data was compiled encompassing 17,218 genes. Principal component analysis (PCA) was applied to identify and exclude gene expression outliers beyond a 95% confidence interval from the grand mean. Although no significant outliers in gene expression were detected, two anomalous samples—from the caudate and DLPFC—were discovered as misaligned during PCA clustering with their respective anatomical regions. To maintain the robustness of the data, these samples were excluded from all further analyses.

#### Replication cohort

A final normalized data was compiled encompassing 18,601 genes. Similarly, PCA was applied to identify and exclude gene expression outliers beyond a 95% confidence interval from the grand mean. A total of 16 samples were identified as outliers according to this metric, and these samples were excluded from all further analyses. This stringent approach ensures the robustness of the dataset and enhances the reliability of downstream analyses. The resulting final dataset included 38 ED cases (34 technical replicates and 4 unique biological replicates), 36 neurotypical controls (26 technical replicates and 10 unique biological replicates), and 37 MDD non-ED neuropsychiatric controls.

### Cellular deconvolution of bulk tissues

We applied a cellular deconvolution algorithm to our bulk RNA-seq data to discern shifts in cellular composition, using the non-negative least squares (NNLS) approach from the MIND R package^33^ with the Darmanis et al., 2015^34^ signature matrix. This matrix comprises six primary cell types: mature excitatory neurons, mature inhibitory neurons, astrocytes, oligodendrocytes, and microglia. The NNLS method, executed via the est_frac function in the *limma* package^35^, was employed on log2-transformed counts per million (CPM) data. Our analysis targeted these principal cell types to minimize variability and to reflect cell type distributions corroborated by preceding research by us and others.

### Computing gene expression variance explained by technical and biological factors

We utilized the variancePartition R package^36^ to construct a linear mixed model (LMM) that would be used to guide downstream differential gene expression testing. The LMM quantified the influence of several technical, biological, and clinical factors on gene expression variability. For each gene within the respective brain regions, variability was ascribed to factors such as estimated neuronal proportions, percentage of intronic reads and rRNA, RNA integrity number (RIN), biological age and sex, library depth, clinical diagnosis, and ethnicity. Variance not accounted for by these factors was catalogued as residual variability.

### Differential gene expression

Differential gene expression analysis was conducted using the *limma* package^35^. We first performed differential expression analysis between the DLPFC and caudate, given their distinct gene profiles revealed through PCA. For this analysis, we used a VOOM normalized matrix across all samples which were not identified as outliers. Here, differential gene expression was adjusted for the possible influence of clinical diagnosis, percentage of intronic reads and rRNA reads, sex, age, and RIN. Donor as a repeated measure was controlled for using the duplicateCorrelation function in *limma*. Given the profound differences in gene expression between these two regions, we next performed differential expression analysis within each region to reveal ED and OCD effects. For these analyses, we used two gene matrices which were VOOM normalized within each region. These analyses covaried for the estimated neuronal proportions (see Methods below), percentage of intronic reads and rRNA reads, sex, age, and RIN. Differentially expressed genes (DEGs) for each comparison were considered significant if they passed a Benjamini-Hochberg (BH) adjusted *p*-value threshold of <0.01. A strict threshold of FDR < 1% was used to protect against potential false positive findings.

### Sensitivity analyses and qSVA

Within each brain region, we performed sensitivity analyses for the following observed covariates sequentially: body mass index (BMI), comorbid major depressive disorder (MDD) and substance abuse defined as opioid use and/or alcoholism and/or polysubstance abuse and/or cannabis use. For each sensitivity analysis - which considered a single confounder from the previous list, we adjusted the original model to include each additional covariate, and then compared the original log2 fold-changes for each diagnosis to the further-adjusted diagnosis coefficient. We also applied the qSVA framework for RNA quality correction in differential expression analysis^37^. This approach aims to reduce the number of false positive genes which may be impacted by RNA quality differences. Quality surrogate variables (qSVs) were estimated using the qsva() function implemented in the sva package^38^. The pairwise Pearson’s correlation between five qSVs, known covariates were investigated using the cor_mat() R function. For comparative case-control analyses in the discovery cohort, within the DLPFC and caudate, we incorporated 15 and 14 qSVs as covariates, respectively.

### Weighted gene co-expression network analysis

Weighted gene co-expression network analysis (WGCNA)^39^ was used to build two signed co-expression networks using a total of 18,486 genes in the DLPFC and 18,472 genes in the caudate. VOOM normalized gene expression data were used as input. To construct each network, the absolute values of Pearson correlation coefficients were calculated for all possible gene pairs in the DLPFC and caudate, respectively, and resulting values were transformed with an exponential weight (β) so that the final matrices followed an approximate scale-free topology (*R*^2^). Thus, for each network we only considered powers of β that lead to a network satisfying scale-free topology (i.e., *R*^2^ > 0.80), so the mean connectivity is high and the network contains enough information for module detection. A β of 5 was used for the DLPFC and a β of 9 was used for the caudate. The dynamic tree-cut algorithm was used to detect network modules with a minimum module size set to 50 and cut tree height set to 0.999.

After network construction, two follow-up analyses were performed: (1) First, a series of module preservation analyses sought to determine whether underlying gene co-regulatory patterns in the DLPFC were preserved or disrupted when compared to the caudate, and vice versa. For these analyses, module preservation was assessed using a permutation-based preservation statistic, *Z*_summary_, implemented within WGCNA with 500 random permutations of the data^40^. *Z*_summary_takes into account the overlap in module membership as well as the density and connectivity patterns of genes within modules. A *Z*_summary_ score <2 indicates no evidence of preservation, 2<*Z*_summary_<10 implies weak preservation and *Z*_summary_ >10 suggests strong preservation. (2) Second, the identified co-expression modules were inspected for association to ED, OCD and ED+OCD as well as all recorded covariates within each region independently. To do so, singular value decomposition of each module expression matrix was performed and the resulting module eigengene (ME), equivalent to the first principal component, was used to represent the overall expression profiles for each module per sample. Modules were evaluated both quantitatively and qualitatively for expression patterns significantly associated with a clinical diagnosis. For quantitative analyses, we computed module significance scores using ME values and clinical diagnoses were treated as main outcomes using a moderated *t*-test through *limma*. These analyses were conducted to match the modelling and covariate adjustments performed for differential gene expression (described above). For qualitative analyses, we generated gene dendrogram trees for each anatomical region with color bars, indicating gene-to-trait associations. Candidate modules were subjected to protein–protein interactions using the STRING database^41^. STRING implements a scoring scheme to report the confidence level for each direct protein–protein interaction (low confidence: <0.4; medium: 0.4–0.7; high: >0.7). We used a combined STRING score of >0.4. Hub genes within the protein–protein interaction network is defined as genes with the highest degree of network connections.

### Gene ontology and enrichment analysis

Correlation adjusted mean rank (CAMERA) gene set enrichment^42^ was performed using the resulting sets of differential gene expression summary statistics between controls with ED, OCD and ED+OCD. We used CAMERA to perform a competitive gene set rank test to assess whether the genes in each gene set were highly ranked in terms of differential gene expression relative to genes that are not in the gene set. For example, CAMERA first ranks gene expression differences in ED DLPFC tissues relative to controls. Next, CAMERA tests whether the user-defined gene sets are over-represented toward the extreme ends of this ranked list. After adjusting the variance of the resulting gene set test statistic by a variance inflation factor that depends on the gene-wise correlation (which we set to default parameters, 0.01) and the size of the set, a *p*-value is returned and adjusted for multiple testing. We used this function to test for enrichment of gene sets derived from Gene Ontology (GO) biological process, molecular factors and genes known to be dynamically regulated throughout human brain prenatal and postnatal brain development. To annotate co-expression modules, genes for each candidate module were used as input for ToppGene^43^ to test for over-representation for GO: Molecular Functions, GO: Biological Processes, and GO: Cellular Components. SynGo^44^ annotation was also used to describe potential enrichment for synaptic terms.

### Secondary analyses of existing OCD transcriptomic datasets

In our secondary analyses, we interrogated 42 raw fastq RNA-sequencing files sourced from Lisboa et al., 2019, comprising data from individuals with OCD (*N*=6) and controls (*N*=8) sampled across three specific brain regions: the putamen, nucleus accumbens, and caudate. Rigorous data processing and quality control measures were employed, ensuring normalization and testing for differential gene expression were consistent with the methodologies detailed above to reduce technical variance. Adjustments were made in the differential expression analysis to account for potential confounding factors of estimated proportions of neuronal populations, intronic read coverage, sex, and biological age. Complementing this, we incorporated differential gene expression summary statistics from Piantadosi et al., 2021, involving a comparison between individuals with OCD (*N*=7) and controls (*N*=8) that focused on the caudate and accumbens. Due to the unavailability of raw RNA-sequencing data, we relied on the summary statistics provided. Lastly, our analysis leveraged differential gene expression data from Jaffe et al., 2014, derived via Illumina HumanHT-12 v3 microarrays from the prefrontal cortex of individuals with eating disorders (ED) (*N*=15), OCD (*N*=16), and a control cohort (*N*=102).

A meta-analysis was conducted utilizing the restricted maximum likelihood (REML) method from the metafor R package, applying the log 2-fold-change alongside standard errors obtained from three distinct OCD comparative transcriptome-wide RNA-sequencing datasets focused on the caudate. These datasets included the current investigation with 27 OCD cases, supplemented by data from Lisboa et al., 2019 (comprising 6 OCD cases), and Piantadosi et al., 2021 (encompassing 7 OCD cases). The analysis was inclusive of genes that were consistently detected across all participant groups, totaling 13,004 genes. To mitigate the risk of false discoveries, the Benjamini–Hochberg (BH) procedure was implemented to adjust the false discovery rate (FDR). Genes were subsequently classified as significantly associated with OCD if they met the BH-adjusted FDR threshold of *p <* 0.01.

### Transcriptome-wide association study (TWAS)

TWAS is a genetic approach that enhances traditional differential expression analyses by integrating estimates of genetically regulated mRNA expression with genetic variant effects on a given phenotype or disorder. Unlike traditional differential expression analyses, which directly measure mRNA levels in cases and controls, TWAS leverages genetically predicted expression by modeling how genetic variants influence mRNA expression. This enables the identification of genes whose predicted expression is associated with disease risk, assigning a directionality (e.g., upregulation or downregulation) based on the estimated genetic effect. In this study, we applied TWAS to prioritize genes potentially involved in ED and OCD. For ED, we used genome-wide significant loci reported from a large-scale GWAS (16,992 cases and 55,525 controls). While no genome-wide significant SNPs have been identified for OCD, we utilized GWAS summary statistics from a previous analysis of 33,943 individuals from the general population. The GWAS summary statistics for both disorders were converted from hg19 to hg38 genome coordinates to ensure consistency. We harmonized SNP names and coordinates across our sample datasets and the GWAS summary statistics. VOOM normalized gene expression matrices were used as input, including18,486 genes from the DLPFC discovery cohort, 18,472 genes from the caudate discovery cohort, and 18,601 genes from the caudate replication cohort. Using the FUSION TWAS framework developed by Gusev and colleagues (http://gusevlab.org/projects/fusion/), we computed gene-level feature weights for the caudate region and applied these weights to the GWAS summary statistics for both ED and OCD. This allowed us to calculate functional-GWAS association statistics, effectively linking genetically regulated mRNA expression with disease risk. The FUSION method was chosen for its ability to estimate the association between gene expression and phenotype by leveraging cis heritability of gene expression. This approach is particularly useful when direct measurements of expression are unavailable or incomplete, as it infers the contribution of genetically regulated expression to disease risk.

## RESULTS

We generated deep bulk/homogenate RNA sequencing (RNA-seq) data from postmortem human tissue in the dorsolateral prefrontal cortex (DLPFC) and caudate from neurotypical controls (*N*=86) and from individuals with a clinical diagnosis of an ED (*N*=57) or OCD (*N*=27) of diverse ethnicity and biological sex (**Figure 1A**). Of the ED cases, eight were diagnosed with anorexia nervosa restricting type (AN-R), 25 were diagnosed with binging and purging behaviors (AN-BP, *n*=14; bulimia nervosa, *n*=11), and the remaining were unspecified ED. All donors displayed varying MDD and substance abuse comorbidity (**Supplemental Table 1**). Body mass index (BMI) was reduced in the ED group compared to controls (*p*=0.009), but not significantly different from the MDD group (*p*=0.18, Mann-Whitney U). Extensive and rigorous quality control of RNA-seq data explored the main drivers of gene expression variability in the current dataset (**Supplemental Figures 1-3**). Cell type proportions were estimated from the bulk tissues and revealed elevated neuronal content in the DLPFC (∼70.8%) relative to the caudate (∼45.5%) (*p*=1.9×10^-30^; Mann Whitney-U test) (**Supplemental Figure 1**). Principal component analysis clearly distinguished DLPFC from caudate tissues along PC1 (**Supplemental Figure 1C**) with a substantial portion (∼86%) of the measured transcriptome differentially expressed at transcriptome-wide significance (FDR<1%) between these two areas (**Supplemental Figure 2, Supplemental Table 2**). Due to these differences, we opted to analyze these two subregions independently for all downstream analyses.

**Figure 1.**
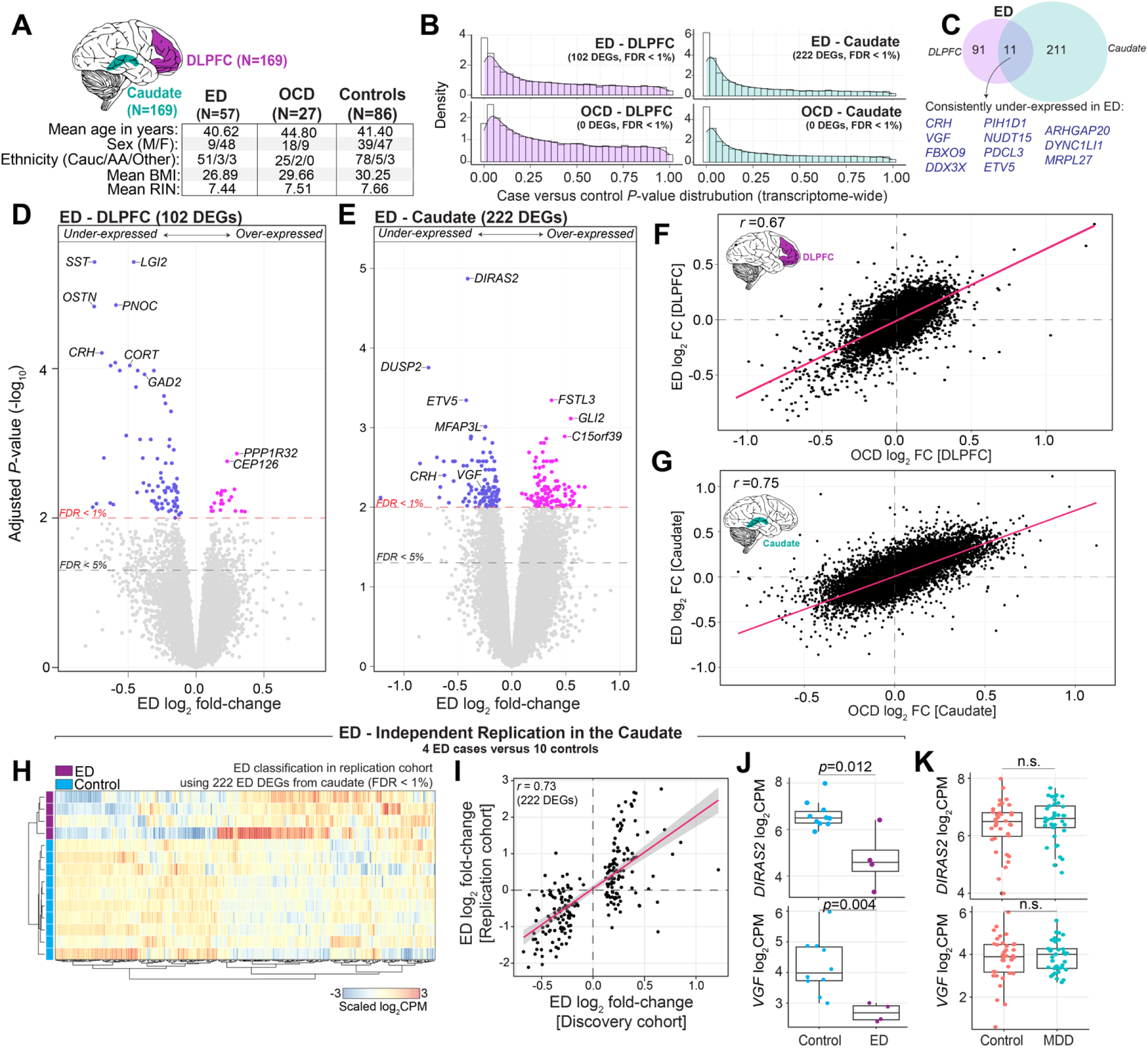
Regional and disorder-specific differential expression analysis. (**A**) **Data Overview**: Bulk RNA sequencing (RNA-seq) data encompasses 169 donors, examining the dorsolateral prefrontal cortex (DLPFC) and caudate nucleus. (**B**) ***P*-value Distribution**: Density plots illustrate an anticonservative *p*-value distributions from differential expression analysis between eating disorder (ED) and obsessive-compulsive disorder (OCD) cases versus controls in the DLPFC (left) and caudate (right). (**C**) **Gene Expression Overlap**: Venn diagrams show the overlap of differentially expressed genes (FDR <1%) for ED in the DLPFC and caudate. (**D-E**) **Volcano Plots**: Depict gene-level effects for ED in the DLPFC (**D**) and caudate (**E**), with the red dotted line marking FDR < 1% and the grey dotted line marking FDR < 5%. (**F-G**) **Transcriptome-wide Concordance**: Scatter plots compare log2 fold-change for ED and OCD gene-level effects in the DLPFC (**F**) and caudate (**G**). Spearman’s correlation coefficients (R-values) are calculated to assess concordance. **(H-J) Replication of ED signatures in the caudate:** (**H**) An independent caudate cohort consisting of 4 ED cases and 11 controls was analyzed to assess the classification ability of the previously identified 222 DEGs in the caudate. Using unsupervised clustering with the Ward distance metric, all 222 genes were collected and used to cluster the independent samples, which are displayed on a heatmap (gene expression is scaled across samples). (**I**) Scatter plots comparing log2 fold-changes of the 222 DEGs between the discovery cohort (x-axis) and the independent replication cohort (y-axis). (**J**) Boxplots validating the expression of top ED genes in the caudate, *DIRAS2* and *VGF*. Multiple test corrected p-values are displayed and were derived from transcriptome-wide moderated *t*-tests. (**K**) Boxplots confirming a null MDD effect for the top ED genes in the caudate.

### Discovery of differential gene expression in ED and OCD

To identify critical covariates for inclusion in our comparative analysis, a linear mixed model quantified the fraction of expression variance attributable to known clinical and technical factors for each gene (**Supplemental Figure 3**). Collectively, these variables explained ∼37.5% and ∼54.8% of transcriptome variation in the DLPFC and caudate, respectively. In the DLPFC, variation in neuronal cell type proportions displayed the largest transcriptome-wide effect that explained a median 6.8% of the observed variation, followed by the percentage of intronic reads (5.7%), percentage of rRNA (5.1%), RIN (1.8%), and biological age (0.47%), and the remaining factors explained a smaller fraction of overall transcriptome variation. Expression variation due to clinical diagnosis (*i.e.* ED and OCD) had a detectable effect in a subset of genes. Notably, equivalent profiles, delineating the relationships between various known factors and gene expression variance, were obtained independently in the caudate (**Supplemental Figure 3C-D**).

We next tested for differentially expressed genes (DEGs) among ED cases versus control donors adjusting for the possible influence of estimated neuronal cell type proportions, RIN, age, sex, ethnicity, and percentage of intronic and rRNA reads. This revealed 102 ED DEGs in the DLPFC and 222 ED DEGs in caudate at strict transcriptome-wide significance (FDR<1%) (**Figure 1B**). Notably, the DEGs exhibited region-specific patterns with minimal overlap (**Figure 1C**) – only 11 genes showed consistent under-expression in both the DLPFC and caudate. We highlight representative DEGs, including decreased expression of Somatostatin (*SST*) and Cortistatin (*CORT*) in the DLPFC, which are expressed in a subpopulation of GABAergic inhibitory neurons, as well as decreased expression of VGF Nerve Growth Factor (*VGF*) and Corticotropin Releasing Hormone (*CRH*) across both the DLPFC and caudate (**Figure 1D-E**), which play crucial roles in neuroendocrine regulation. Sensitivity analysis corroborated the stability of our findings, even when considering differences related to BMI, MDD comorbidity and substance abuse histories (**Supplemental Figure 4**).

No transcriptome-wide significant DEGs (FDR <1%) were detected for OCD in either brain region, which may be a function of a restricted OCD sample size. However, comparative examination of the transcriptome-wide effects between ED and OCD indicated a high degree of concordance across both disorders within each region — DLPFC (R=0.67) and caudate (R=0.75) — despite the absence of significant DEGs for OCD (**Figure 1F-G**). This consistency in gene expression directionality points to a potential molecular intersection between ED and OCD, consistent with shared underlying pathophysiological mechanisms. To substantiate this, we conducted a direct comparative differential gene expression analysis between individuals with ED versus OCD and found no significant DEGs (**Supplemental Figure 5**), reinforcing the notion of shared, rather than disorder-specific, gene dysregulation. Overall, the pronounced gene expression differences in the caudate, coupled with the parallel effects observed in both ED and OCD across these key brain regions, emphasize a molecular convergence indicative of common pathophysiological pathways in these disorders.

### Replication of ED transcriptional signatures in the caudate

Considering the pronounced caudate effect in ED, we sought to replicate these findings within our smaller RNA-seq replication cohort. This cohort focused on the caudate of 38 ED cases (34 technical replicates and 4 unique biological replicates), 36 neurotypical controls (26 technical replicates and 10 unique biological replicates), and 37 MDD non-ED neuropsychiatric controls, all from an all-female cohort of European descent (**Supplemental Table 1**). First, we confirmed the validity our quality control metrics, including estimated cell type proportions, gene expression variability attributable to known factors, and covariate concordance in the caudate (**Supplemental Figure 6**). Next, transcriptome-wide analysis of 34 ED cases and 26 controls, which were technical replicates between the discovery and replication cohorts, accounted for neuronal proportions, RIN, age, sex, and percentage of intronic and rRNA reads. This analysis confirmed a high degree of transcriptome-wide replication for the ED caudate signatures (*r*=0.86) (**Supplemental Figure 7**), underscoring the robustness of these DEGs relative to technical variability.

Notably, we leveraged the remaining unique biological replicates from the replication cohort, comprising a non-overlapping set of 4 ED cases and 10 controls. We tested the classification performance of the 222 ED DEGs derived from the discovery cohort to distinguish these independent ED cases from controls using unsupervised hierarchical clustering **(Figure 1H)**. This analysis accurately classified all ED cases from controls, demonstrating the robustness and predictive power of the ED DEG caudate signatures in a small independent set of samples. The validation of both increased and decreased expression levels of these DEGs between the discovery cohort and these non-overlapping samples was observed (*r*=0.76) (**Figure 1I**). This replication reinforces the dysregulation of top ED genes in the caudate, including top ED genes *DIRAS2* and *VGF* (**Figure 1J**). The consistency of these signatures highlights their potential as reliable biomarkers and/or mechanisms for ED. Moreover, while differences in BMI were observed between groups, no significant associations were found between BMI and gene dysregulation in the caudate, further supporting the specificity of these transcriptomic signatures to ED, independent of BMI differences.

Finally, considering that ED cases display varying comorbid MDD, a unique advantage of this replication cohort is the inclusion and joint RNA-sequencing of a MDD non-ED neuropsychiatric control group. This enabled us to directly examine any overlapping gene dysregulation between ED and MDD in a controlled experimental setting. No DEGs were observed between 37 MDD and 36 controls with an FDR < 1% (**Supplemental Figure 8A-B**), and only one gene (*LINC02199*) met an FDR < 5%. Transcriptome-wide concordance among gene-level effect sizes between ED and MDD within the caudate indicated a low correlation (*r*=0.19) (**Supplemental Figure 8C**), with ED-related genes, including *DIRAS2* and *VGF*, showing no significant changes in MDD (**Figure 1K**). These results complement our initial sensitivity analyses (**Supplemental Figure 4**), supporting that ED DEGs are indeed not confounded by comorbid MDD.

### Combined ED and OCD transcriptional analysis

Considering the positive concordance in transcriptional profiles between ED and OCD (**Figure 1F-G**), we conducted a unified analysis comparing the gene expression profiles of combined ED and OCD cases (ED+OCD) against control groups within each respective brain subregion. The aggregated analysis for ED+OCD uncovered a heightened count of significant DEGs at a stringent false discovery rate (FDR < 1%): 232 in the DLPFC and 815 in the caudate (**Figure 2A**). Notably, a substantial proportion of these DEGs overlapped with those identified in the ED-only analysis (81.3% in the DLPFC and 90% in the caudate). Of these, 56 genes were jointly dysregulated across both regions, including genes involved in neuroendocrine regulation *CRH*, *VGF* and tachykinin precursor 1 (*TAC1*). Importantly, the directional trends of these DEG signatures remained consistent even after correcting for potential RNA degradation effects (qSVA), indicating that the robustness of our results is not attributable to sample quality concerns (**Supplemental Figure 8**). In the DLPFC, *SST* was notably diminished, along with other key genes like corticotropin releasing hormone binding protein (*CRHBP*) and brain derived neurotropic factor (*BDNF*), both essential for neurodevelopment and synaptic function. (**Figure 2B**). We also observed decreased expression of the GABAergic synaptic genes glutamate decarboxylase 1 and 2 (*GAD1*, *GAD2*) and solute carrier family 32 member 1 (*SLC32A1*), genes that are previously suggested to contribute to the pathophysiology of OCD and related disorders^20^. The caudate analysis showed a significant downregulation of genes, including GTP-binding Ras-like protein 2 (*DIRAS2*), implicated in ADHD susceptibility^45^, and dual specificity phosphatase 2 protein (*DUSP2*), associated with neuronal maintenance and various neurological conditions^46,47^ (**Figure 2C**). We employed a competitive gene set ranking method to annotate these DEGs functionally, which showed a significant enrichment in genes involved in mitochondrial translation, oxidative phosphorylation, and RNA stability, underscoring their potential role in both ED and OCD (**Figure 2D-E, Supplemental Table 3**). These under-expressed genes also corresponded to those exhibiting increased expression from prenatal to postnatal stages of cortical development. Furthermore, genes implicated in these downregulated pathways notably interact with Chromodomain Helicase DNA Binding Protein 8 (CHD8)^48^, identified as an OCD-associated risk factor^49–50^, suggesting a common thread in the genetic landscape of these disorders.

**Figure 2.**
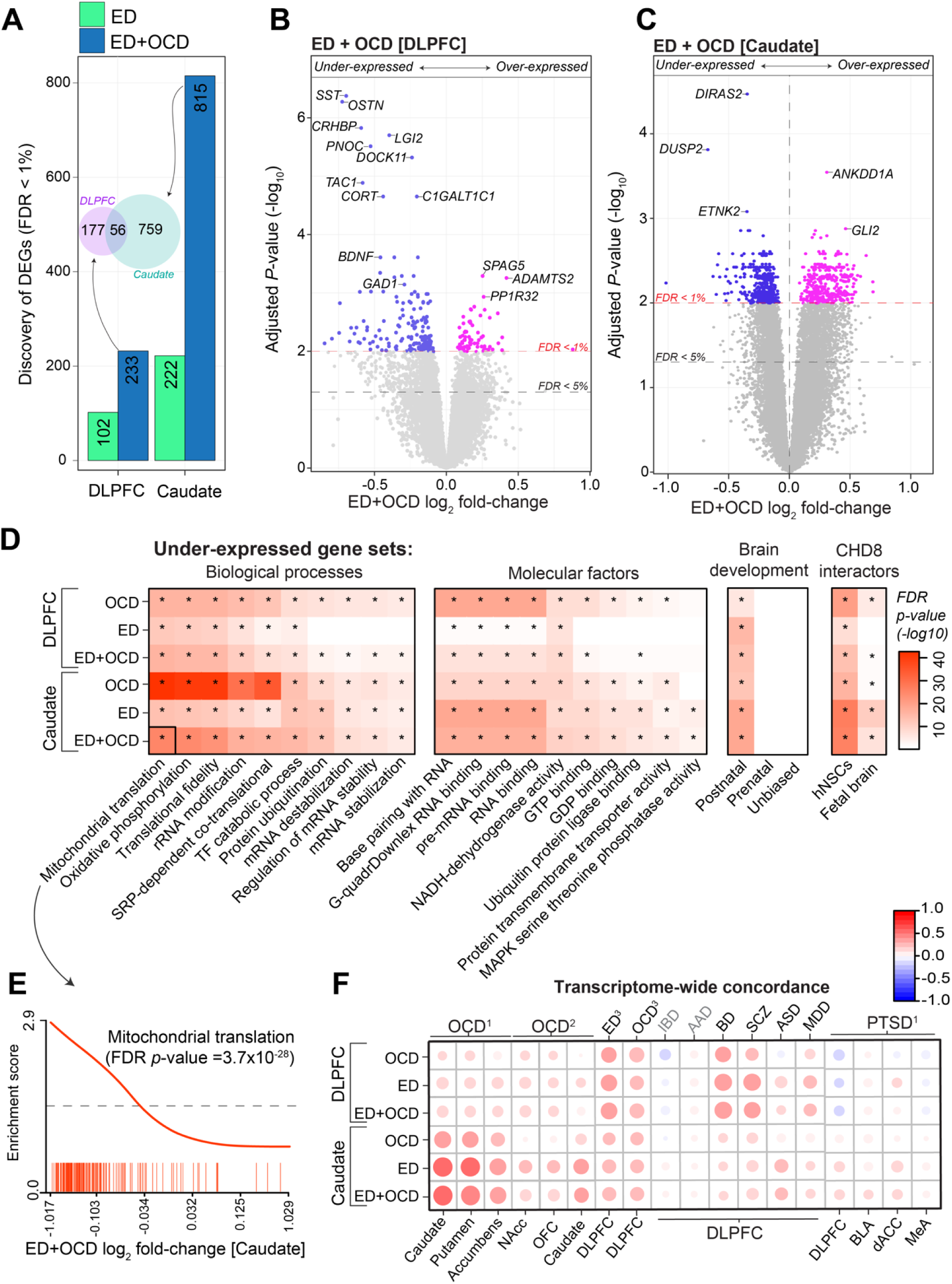
Integrated Analysis of Gene Expression in ED and OCD. (**A**) **Differential Gene Count**: The bar graph displays the count of differentially expressed genes (DEGs) with False Discovery Rate (FDR) < 1% on the y-axis, contrasting controls against cases of ED alone and the combination of ED with OCD (ED+OCD) across both brain regions shown on the x-axis. An inset Venn diagram illustrates the significant ED+OCD DEG overlap between the regions. (**B-C**) **Volcano Plots for Combined Analysis**: Displaying the significance (-log10 FDR p-value; y-axis) versus magnitude of expression change (log2 fold-change; x-axis) for ED+OCD DEGs in the DLPFC (B) and caudate (C). (**D**) **Gene-Set Enrichment Analysis (CAMERA)**: Heatmap showing gene-set enrichment analysis outcomes for genes under-expressed in OCD, ED, and ED+OCD within the caudate and DLPFC (y-axis), with FDR-adjusted p-values for each gene set. Notably enriched categories include mitochondrial translation, oxidative phosphorylation, mRNA stability, genes with postnatal expression bias, and CHD8 interacting proteins. Minimal to no enrichment is observed for over-expressed genes. (**E**) **CAMERA Enrichment Barcode Plot**: An example barcode plot highlights gene-set enrichment for mitochondrial translation on the y-axis, plotted against a rank of ED+OCD-related genes sorted by log2 fold-change on the x-axis. This example corresponds to the black-outlined section of the heatmap in panel D. (**F**) **Transcriptome-wide Concordance**: A scatter plot compares transcriptome-wide gene-level effect sizes for OCD, ED, and ED+OCD alterations against those derived from independent brain transcriptome studies (y-axis), demonstrating high correlations, particularly with independent studies of OCD. References: OCD^1^ (Lisboa et al., 2019), OCD^2^ (Piantadosi et al., 2021), ED^3^OCD^3^ (Jaffe et al., 2014) and PTSD^1^ (Jaffe et al., 2022).

### Concordance with transcriptional analyses in independent samples

To contextualize our findings within the broader landscape of neuropsychiatric research, we aggregated transcriptome-wide summary statistics from prior bulk postmortem brain tissue RNA-seq studies, encompassing nine distinct disorders (**Supplemental Table 4**). This analysis was intended to evaluate the congruence between our transcriptome-wide log_2_ fold change signatures and those documented in various clinically related but distinct disorders. Our analysis yielded two key insights (**Figure 2F**). Primarily, the transcriptomic patterns we identified for ED, OCD, and the combined ED+OCD within the caudate showed strong replication and the highest alignment with transcriptomic data from separate smaller OCD and ED studies. Specifically, correlations with OCD-related transcriptome changes from Lisboa et al., 2019^19^ (*n*=6 OCD cases), showed substantial agreement in the caudate, putamen and accumbens with our ED (*R*=0.56, *R*=0.52, *R*=0.39), OCD (*R*=0.42, *R*=0.40, *R*=0.37), and combined ED+OCD (*R*=0.60, *R*=0.57, *R*=0.43) signatures (**Supplemental Figure 9**). A similar pattern of high correlation was evident with OCD-related findings from Piantadosi et al., 2021^21^ (*n*=7 OCD cases), in the nucleus accumbens, orbitofrontal cortex (OFC), and caudate relative to our ED (*R*=0.37, *R*=0.38, *R*=0.42, respectively) and ED+OCD (*R*=0.32, *R*=0.35, *R*=0.40, respectively) signatures in the caudate. We also observed high transcriptome-wide correlations with ED-related findings from Jaffe et al., 2014^22^ (*n*=15 ED cases) in the DLPFC with our ED (*R*=0.35), OCD (*R*=0.31) and ED+OCD (*R*=0.36) signatures in the DLPFC.

Furthermore, our analysis indicated that independent bipolar disorder (BD) and schizophrenia (SCZ) signatures from the DLPFC^51^ were more closely aligned with DLPFC signatures for ED, OCD, and combined ED+OCD rather than those from the caudate, suggesting a regional specificity in gene expression patterns. Additionally, the minimal or absent overlap of alcohol abuse disorder (AAD) and inflammatory bowel disorder (IBD) signatures^51^ with our data implies that the transcriptomic similarities we observed are unlikely to be attributed to comorbid substance abuse, overall health status, or generic post-mortem brain changes, and bolster the validity of our findings.

### Meta-analyzing OCD transcriptional signatures from the caudate

Given the convergence of transcriptome-wide OCD gene expression signatures within the caudate and acknowledging the unique transcriptional distinctions between the caudate and DLPFC, we advanced our findings through a gene-based meta-analysis. This step was taken to augment the sample sizes for OCD and amplify the identification of genes dysregulated in OCD by integrating results from extant studies focusing on the OCD caudate transcriptome. This meta-analysis leveraged a combined sample size from three studies, encompassing the caudate from 40 individuals with OCD: our current cohort (N=27), alongside those from Lisboa et al., 2019 (N=6) and Piantadosi et al., 2021 (N=7). Our meta-analysis revealed 57 genes exhibiting significant differential expression in OCD with an FDR < 1% (**Figure 3, Supplemental Table 5**), signaling robust alterations in the caudate. Among these, we observed under-expressed *DIRAS2* and the over-expressed GLI Family Zinc Finger 2 (*GLI2*), both of which were prominent and significantly dysregulated in ED, reinforcing the joint ED+OCD analysis findings. We also found over-expression of several cytokine receptor activity genes in OCD caudate, including interleukin 10 receptor subunit beta (*IL10RB*), interleukin 11 receptor subunit alpha (*IL11RA*) and interferon lambda receptor 1 (*IFNLR1*).

**Figure 3.**
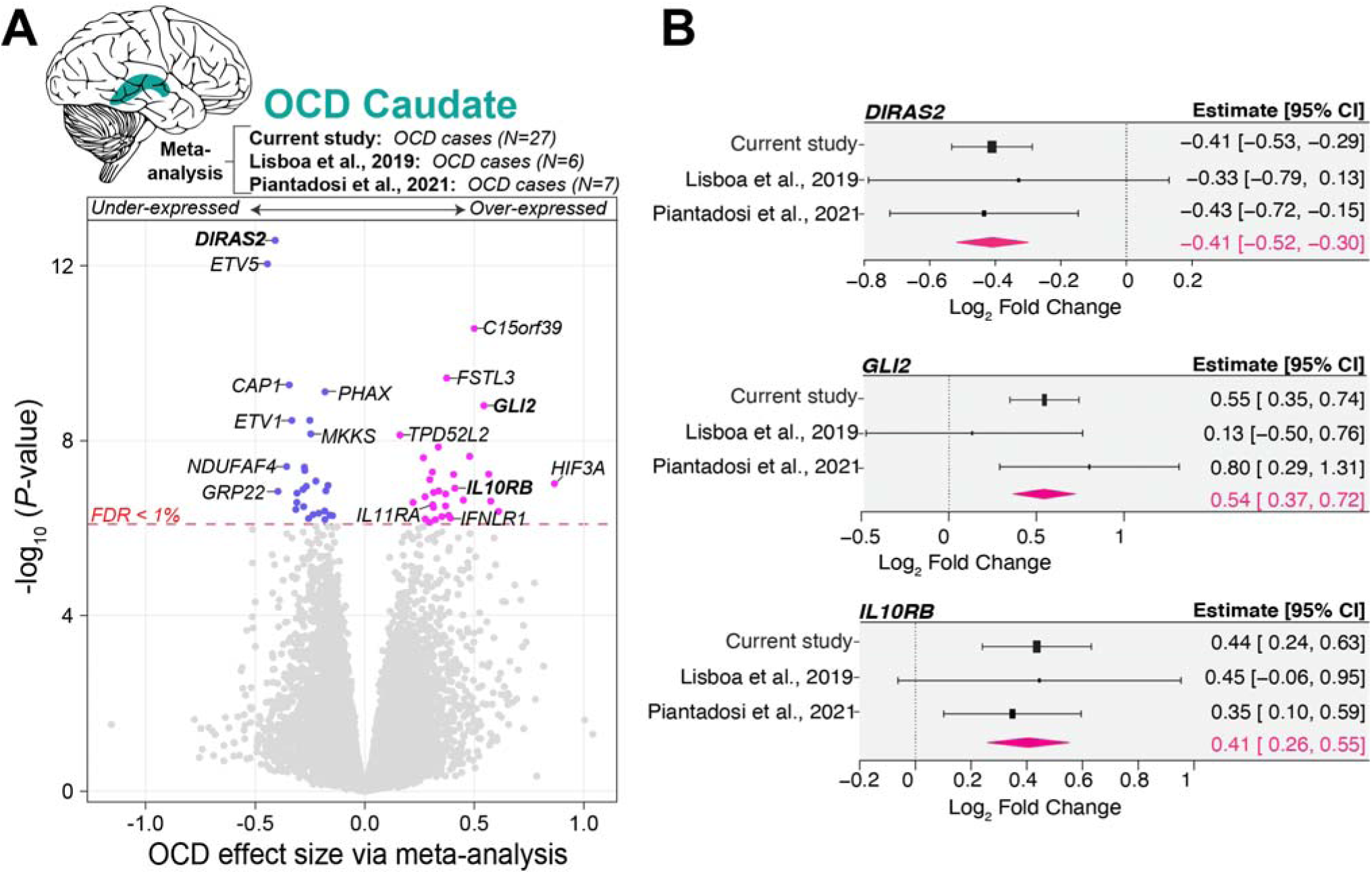
Meta-analysis of OCD Gene Expression Signature in the Caudate. (**A**) A volcano plot that synthesizes the differential gene expression (DEG) signatures associated with OCD in the caudate nucleus. The plot juxtaposes the significance of gene expression changes (-log10 p-value; y-axis) against the magnitude of these changes (effect size [beta]; x-axis), aggregating data from 40 OCD cases across three distinct studies. Our meta-analysis reveals that, within the caudate of individuals with OCD, 30 genes are significantly over-expressed, and 27 genes are under-expressed, achieving an FDR threshold of less than 1%. The integration of DEG summary statistics was conducted using the metafor package in R. Gene symbols in bold are highlighted in panel B. (**B**) Forest plots highlight meta-analysis of log_2_ fold change (log_2_FC) in gene expression for the specified gene across three independent studies. The x-axis shows the log fold change, with each plot’s x-axis dynamically set to the minimum and maximum confidence interval (CI) values specific to that gene, allowing for accurate visualization of effect sizes. For each gene, individual studies are listed along the y-axis, with markers indicating the logFC for each study and error bars representing the 95% confidence intervals. The summary diamond (pink) at the bottom of each plot reflects the combined random-effects model estimate, summarizing the overall effect size across studies.

### Gene co-expression network analysis

To refine our understanding of the biological pathways and cellular mechanisms involved, we extended our investigation through weighted gene co-expression network analysis (WGCNA). This approach allowed us to pinpoint discrete groups of co-expressed genes, termed modules, within the DLPFC and caudate, respectively. A total of 13 modules were identified in the DLPFC and 15 modules in the caudate (**Supplemental Figure 10**). The integrity of these co-expression modules was largely preserved across both brain regions, with most variances observed in the smaller modules (**Supplemental Figure 10**).

Particularly noteworthy were two modules, Md1 in the DLPFC and Mc1 in the caudate. These modules demonstrated a clear neuronal profile (**Figure 4A-B, Supplemental Figure 11**), enriched for DEGs, and were consistently down-regulated in ED+OCD (**Figure 4C**). These modules stood out due to their enrichment for proteins that interact with CHD8, a gene associated with OCD risk (**Figure 4D**) and their significant role in mitochondrial translation and oxidative phosphorylation pathways (**Figure 4D**), both vital to cellular energy and neuronal function. They also shared a functional enrichment for pre-synaptic vesicle activities (**Figure 4E**), which are crucial for neurotransmission. A striking 76.1% of the genes in the caudate’s M1c were also present in DLPFC’s M1d. The depth of shared protein-protein interactions within these modules further supports the complex interplays that could be fundamental to understanding the neural and synaptic dysfunctions shared across these disorders (**Supplemental Figure 12**).

**Figure 4.**
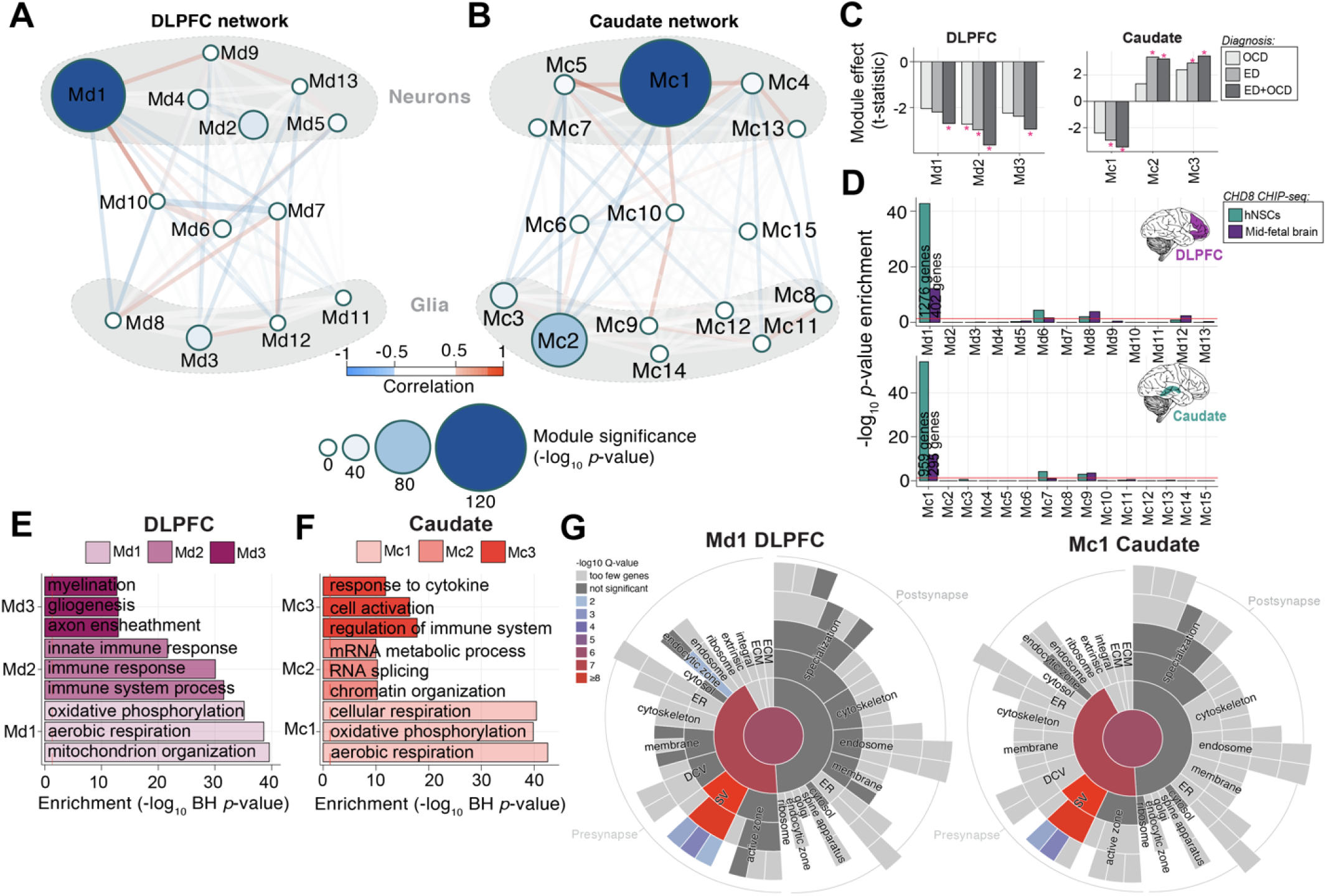
Network analysis identifies modules of co-expressed genes across ED and OCD. (**A-B**) **Module Relationships and Cell Type Clustering**: Network diagrams illustrate the relationships between gene co-expression modules in the DLPFC (A) and caudate (B), with module size representing the -log10 p-value significance of ED+OCD DEG enrichment in each region. (**C**) **Module-Level Differential Expression**: Bar graphs depict t-statistics from a moderated t-test model, indicating module eigengene association with disease status (FDR-corrected *P<0.05). (**D**) **CHD8 Interacting Proteins Module Enrichment**: Enrichment levels (-log10 p-value, Fisher’s Exact Test) for CHD8 interacting proteins across modules are shown for human neural stem cells (hNSCs) and human mid-fetal brain tissue, with separate analyses for DLPFC modules (top) and caudate modules (bottom). (**E-F**) **Functional Enrichment of Top Modules**: The functional enrichment (-log10 Benjamini-Hochberg adjusted P-value; x-axis) for the top three co-expression modules is compared in the DLPFC (E) and caudate (F). (**G**) **SynGO Sunburst Plots for Synaptic Gene-Set Enrichment**: Sunburst plots from SynGO analysis depict the synaptic gene-set enrichment for genes within the most significant modules, Md1 in the DLPFC (left) and Mc1 in the caudate (right).

Additional modules also revealed novel insights. In the DLPFC, module Md2 was down-regulated across all conditions (**Figure 4C**) and notable for its significant concentration of genes related to innate immune responses (**Figure 4E**). Similarly, the Md3 module, characterized by a glial cell signature (**Figure 4A, Supplemental Figure 11**) indicated a down-regulation in ED+OCD (**Figure 3C**) and is implicated in myelination and glial cell development (**Figure 4E**), processes critical for neuroplasticity and brain homeostasis. In the caudate, the Mc2 module was distinctly up-regulated in ED and combined ED+OCD but not extensively in OCD (**Figure 4C**), suggesting disorder-specific gene sets involved in glial cell function, chromatin remodeling and RNA processing (**Figure 4E**). The Mc3 module, which was also up-regulated and glial-specific, focused on genes linked to cellular activation and cytokine response pathways (**Figure 4E**), reinforcing the potential role of glial cells in the pathophysiology of ED and possibly OCD.

Collectively, these findings underscore the roles of modules M1d and M1c in elucidating the common neuropathology of ED and OCD, while also highlighting additional, distinct pathways that could uniquely influence the molecular underpinnings of each disorder, offering a more granular understanding of their differential impacts on brain function.

### Genetically predicted gene expression changes

Transcriptome-Wide Association Study (TWAS) was conducted to complement traditional differential expression analyses by integrating genetically predicted expression levels. TWAS was performed using models of genetically predicted mRNA expression across our discovery cohorts (DLPFC and caudate) and a replication cohort (caudate). The analysis assessed the association between genetically predicted mRNA expression and the odds of ED and OCD, separately. After correcting for multiple testing across all genetic models (18,486 genes in the DLPFC discovery cohort, 18,472 genes in the caudate discovery cohort, and 18,601 genes in the caudate replication cohort), we identified 62 significant association signals for ED, representing 50 unique genes, with several genes replicating across both brain regions (**Supplemental Table 6**). These 50 unique genes were enriched for metabolism-related genes (FDR *p*=0.03), including *IMPDH2*, *SLC25A20*, *GBE1*, *KYNU*, and *DPYD*, Notably, the strongest associations were in a gene-dense region on chromosome 3 (**Supplemental Figure 12**), consistent with previous TWAS reports on ED. This cluster overlaps with a known GWAS signal for Anorexia Nervosa (AN) (chromosome 3: 47,588,253–51,368,253), further reinforcing its relevance. We highlight nine significant genes that replicated across two brain regions and/or cohorts (**Table 1**). Among these, upregulation of WD Repeat Domain 6 (*WDR6*) was one of the top hits, showing a robust association with increased odds of ED (Z = 7.11, p = 1.1 × 10 ¹², DLPFC; Z = 5.54, p = 3.0 × 10, caudate discovery; Z = 7.22, p = 5.3 × 10 ¹³, caudate replication). Similarly, downregulation of NCK Interacting Protein with SH3 Domain (*NCKIPSD*) was also associated with increased ED risk across brain regions. Other notable downregulated genes in the chromosome 3 cluster include *DALRD3*, *C3orf62*, *P4HTM*, and *CTNNB1*, as well as *MGMT* on chromosome 10, were found to increase ED risk. Importantly, many of these genes, including *WDR6* and *NCKIPSD*, have been reported in previous AN TWAS analyses, confirming the robustness of our findings. Additionally, we identified novel associations, such as the downregulation of *LINC02055*, a lincRNA, linked to ED risk in the caudate (Z = -4.23, p = 2.0 × 10 ², caudate; Z = -4.55, p = 6.4 × 10 ³, caudate replication). Regarding OCD, no significant signals were identified likely due to the lack of current GWAS statistical power. We hypothesize that signals not replicated in the current analysis may be due to the FUSION method only utilizing cis-heritable genes or differences in the genetically regulated gene expression models used across methods.

**Table 1.**
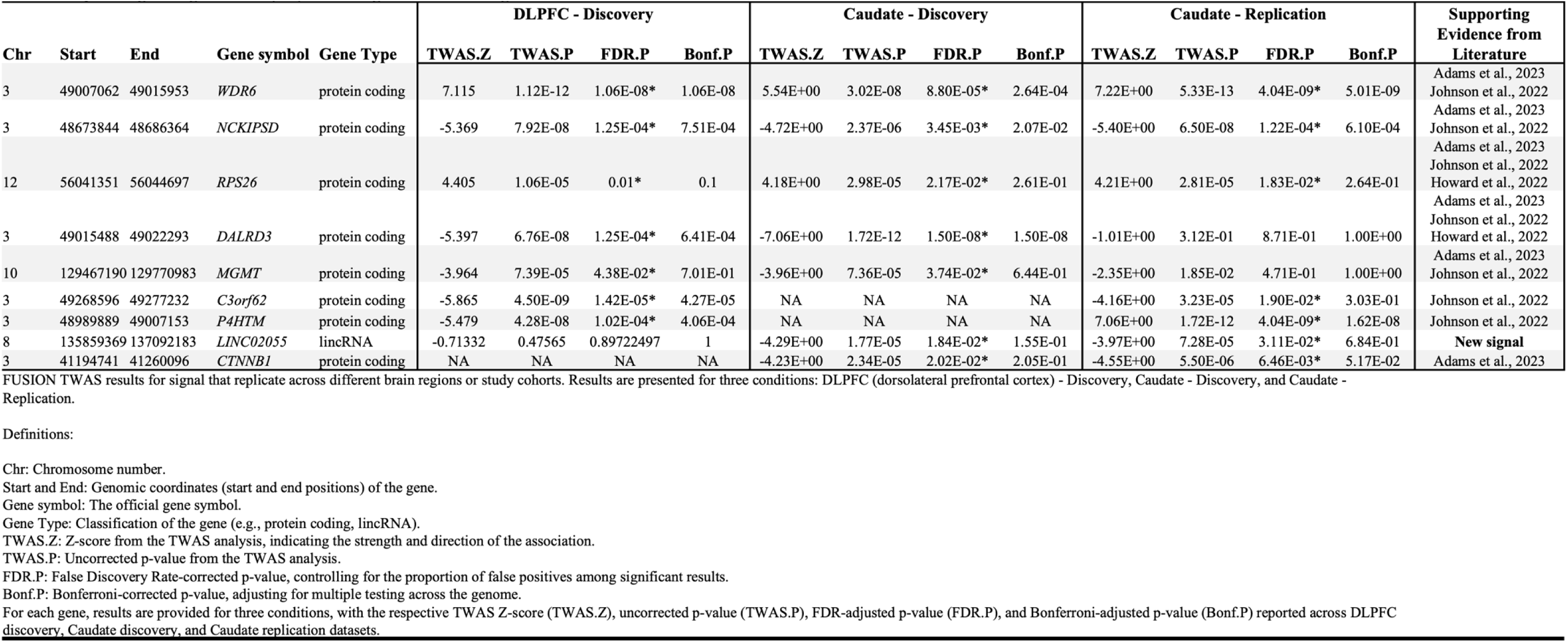
Renlicatim, Eatinl! Disorder fED) TWAS silmal across two rel!ions and/or cohorts.

## DISCUSSION

As our understanding of the clinical manifestations and genetic underpinnings of OCD and ED advances, decoding the functional pathways underlying these disorders has remained challenging. Our study takes a rigorous approach to dissecting the transcriptomic landscapes of two neural subregions central to these disorders

—the DLPFC and caudate. We found significant convergence of gene expression differences, underscoring a molecular link between ED and OCD. The joint analysis of ED and OCD uncovered many down-regulated genes, including those linked to GABAergic neurons and neuroendocrine dysfunction. Additional downregulation of genes associated with cellular processes, including mitochondrial translation, oxidative phosphorylation, and mRNA stability, was observed. Notably, ED caudate signatures were successfully replicated in an independent cohort, and these findings were not likely confounded by MDD comorbidity. These genes present significant insights, particularly through their interactions with CHD8. Genetically regulated gene expression alterations also point towards dysregulated metabolism in ED risk, including top genes *WDR6*, *KCKIPSD*, and *RPS26*. This work thus contributes to a more nuanced understanding of the transcriptomic basis of these disorders and lays the groundwork for decoding the molecular neuropathologies and future development of therapeutic approaches.

The transcriptomic concordance observed between OCD and ED signal a linked molecular landscape, suggesting shared underlying mechanisms. The consistent downregulation of CORT mRNA within the DLPFC and caudate across individuals diagnosed with ED (**Figure 1-2**) reflects potential broader neurobiological implications that transcend the canonical roles in emotion and behavioral regulation. Cortistatin, a neuropeptide encoded by CORT, is recognized for its role in modulating GABAergic neurons^52,53^. Animal studies have shed light on deficits in cortistatin’s pathway leading to neurological anomalies^54,55^. In alignment with these findings, our study reveals significant reductions in a suite of neuroendocrine genes, including *VGF*, *SST*, *TAC1,* and *CRHBP* as well as additional GABAergic markers *GAD1*, *GAD2*, and *SLC32A1.* Notably, this result is not explained by reduced proportions of inhibitory neurons in ED and OCD (**Supplemental Figure 1B**). Previous studies have also indicated a decrease in *GAD1* and *GAD2* expression in OCD^20^, with *GAD2* also identified as a common risk factor for OCD^56^. Additionally, diminished expression of *GAD1* in the striatum of those with Tourette Syndrome highlights the potential shared neuropathological mechanisms^57^. Moreover, both *SST* and *VGF* are inducible by *BDNF* – which itself exhibited reduced expression levels in our study – and synergistically modulates aspects of neuronal activity, viability, neurogenesis, energy balance, lipolysis, and behavior^58,59^. These observations lend support to the hypothesis that anomalies in GABAergic signaling, alongside neuroendocrine system disruptions, are implicated in the shared pathophysiology characteristic of both ED and OCD. These functional domains, integral to the regulation of compulsive behaviors and stress responses^60,61^, are further substantiated by genetic links in anorexia nervosa and related metabolic and immunological disorders^62–64^. This suggests the existence of a common genetic thread predisposing to a spectrum of psychiatric conditions.

Our work also revealed under-expression of *DIRAS2* in OCD and ED in the caudate (**Figure 1,3**). Notably, DIRAS2 has been consistently implicated in ADHD, impaired cognitive control and impulsivity through genome-wide association and genetic linkage studies^45,65–69^, underscoring a broader relevance across distinct but potentially related clinical phenotypes. Located in chromosomal region 9q32.2, the relevance of DIRAS2 is magnified by its function within the Ras superfamily of GTPases^70^—proteins known for their involvement in cellular signaling cascades that regulate critical processes such as cell division and differentiation. Moreover, DIRAS2 has been linked to executive functions through its expression in neural substrates such as the prefrontal cortex, amygdala, and anterior cingulate cortex^69,71,72^ - areas instrumental in executive decision-making and the modulation of anxiety. Recently, significant associations have emerged linking rare variants in DIRAS2 to suicide attempt behaviors in combat survivors^72^, consistent with this gene’s relevance to psychiatric presentations. However, despite its distinct biochemistry within the Ras protein family, the full scope of molecular pathways and cellular functions modulated by *DIRAS2* remains elusive. Unraveling *DIRAS2* molecular functions and pathways represents a critical step for future research. In addition to *DIRAS2*, it is worth noting that dysregulation of multiple other genes in the caudate were more pronounced than in the DLPFC (**Figure 2A**), which may be linked to the caudate nucleus’s role as a component of the basal ganglia – a region involved in procedural learning, associative learning, and habit formation. The caudate has also been implicated in the reward system and the processing of emotions, which are critical elements in both ED and OCD.

The significant decrease in the M1d and M1c modules (**Figure 4**), which are instrumental for the integrity of neuronal function, particularly in protein synthesis and mitochondrial energy production, further underscore a shared molecular dysfunction. The perturbation in these modules suggests a molecular deficit that could compromise synaptic efficacy and plasticity^73,74^. The resulting energy deficiency in neurons is proposed to impede the proper regulation of synaptic transmission^73,74^. This finding may reflect a mechanistic link where insufficient nutrient availability leads to compromised cellular energy synthesis, highlighting the physiological consequences of malnutrition in ED on metabolic processes. These modules were also enriched for downregulation of genes that interact with CHD8, known for its vital role in transcriptional regulation and chromatin remodeling during early neurodevelopment^48^. As the genetics of ED and OCD mature, the discovery of transcriptomic alterations linked to CHD8, a current leading genetic risk variant for OCD^50^, suggests an early promising connection. Targets of CHD8 have also been implicated in bipolar disorder^75^. Notably, OCD is associated with a significantly increased risk of a concurrent or subsequent diagnosis of bipolar disorder^76,77^, suggesting a convergent pathogenic pathway via CHD8 targets across these disorders. Our identification of these transcriptomic changes provides evidence that the genetic and gene expression profiles converge. This is further substantiated by our findings of *TUSC2* down-regulation in ED, a top candidate gene linked to anorexia nervosa through transcriptomic imputation^62^, supporting a genetic and transcriptomic nexus. The identification of such targets underlines the complex molecular landscape shaping these disorders.

Our network approach also highlights the importance of immune response and glial cell functions in the pathophysiology of ED and OCD. The down-regulation of module Md2 in the DLPFC, enriched in genes associated with innate immune responses, aligns with long-standing evidence suggesting the involvement of immune dysregulation in OCD^19,20^. Similarly, the down-regulation of the Md3 module, indicative of altered myelination and glial cell development, supports the hypothesis that neuroplasticity disruptions are central to ED and OCD. These findings reinforce the concept of glial cells as key players in the pathophysiology of these disorders, potentially offering new targets for therapeutic intervention.

The application of TWAS in this study provides key insights that complement traditional differential expression analyses. This approach not only prioritizes disease-relevant genes but also assigns directionality to their expression changes, adding a layer of mechanistic insight. Upregulation of *WDR6* emerged as one of the most significant association signals across brain regions, a finding supported by previous research^78,79^. *WDR6*, a conserved gene involved in protein complex assembly and potentially modulating insulin signaling in the brain. Its repeated association across methods—TWAS, conditional analysis, and fine-mapping—strongly suggests a causal role for *WDR6* in ED. This aligns with existing evidence that implicates insulin dysregulation in ED, further highlighting the potential role of metabolic pathways in the disorder, as reported via transcriptome-wide profiles. Furthermore, several other genes located in the chromosome 3 gene cluster—including *NCKIPSD, DALRD3, C3orf62, P4HTM*, and *CTNNB1*—showed significant downregulation, contributing to the overall risk of ED. These genes have also been identified in prior studies^78–80^, reinforcing their relevance to ED pathobiology. Notably, our study also identifies *LINC02055*, a lincRNA, as a novel signal associated with ED. While the specific function of *LINC02055* in the brain is still unclear, its classification as a long non-coding RNA and its identification in transcriptome studies linked to brain disorders like ED suggest it could play a regulatory role in the genetic and molecular networks that underpin brain function and pathology. Further research will help clarify its potential contributions to neurodevelopmental and psychiatric conditions. Finally, while TWAS provides powerful insights, its limitations, such as potential confounding from linkage disequilibrium and the incomplete representation of gene expression in certain tissues, highlight the need for continued refinement. Larger GWAS datasets, alongside cell-type-specific expression models and ancestry-diverse cohorts, will be crucial for improving the precision of these findings and uncovering additional causal genes.

This study also has limitations inherent to postmortem research, including potential confounding factors such as medication use, substance use, and psychiatric comorbidities. While we conducted extensive sensitivity analyses to account for these variables, it is possible that they may still impact the observed transcriptomic changes. Additionally, while the ED and OCD sample sizes are relatively small when compared to the scope of postmortem brain research for other neuropsychiatric disorders (*e.g.* schizophrenia), the current data represent the largest transcriptomic study of these disorders conducted to date, underscoring the rarity of these cases in medical examiner cohorts. The heterogeneity within the ED sample, encompassing diverse subtypes and clinical presentations, remains a challenge for generalizability. Future work should focus on increasing cohort size and diversity, incorporating cell-type-specific transcriptomic analyses, and investigating alternative models such as peripheral biomarkers or iPSCs to complement findings from postmortem brain tissue.

## CONCLUSIONS

In conclusion, this study sheds new light on the transcriptomic underpinnings of ED and OCD, revealing shared molecular alterations that bear significance for the neuropathology these disorders. We identify reduced expression of genes associated with GABAergic neuron functionality and neuroendocrine regulation, further implicating deficits in energy generation and oxidative phosphorylation. Additionally, we identified links to the OCD-related CHD8 locus, enhancing our understanding of the shared cellular and molecular dysfunctions in these disorders. By juxtaposing our transcriptomic data with findings from other neuropsychiatric disorders, we underscore the broader applicability and relevance of our results. Through our TWAS analysis, we identified key genes, such as *WDR6* and *NCKIPSD*, that are strongly associated with ED, suggesting a critical role for genetically regulated expression in ED pathogenesis. This signal further supports the involvement of dysregulated metabolic and neuroendocrine pathways, potentially modulating energy regulation and synaptic plasticity. These insights not only expand the scope of current ED and OCD research, but also support the development of targeted therapies aimed at correcting these underlying molecular aberrations, ultimately steering us toward more effective clinical management.

## Data Availability

All original RNA-sequencing data are publicly at the National Center for Biotechnology Information Gene Expression Omnibus under the following accession number - GSE262138.

https://www.ncbi.nlm.nih.gov/geo/

## Author Contributions

Dr. Breen had full access to all the data in the study and takes responsibility for the integrity of the data and the accuracy of the data analysis.

*Concept and design*: All authors.

*Acquisition, analysis, or interpretation of data*: MSB and all authors.

*Drafting of the manuscript*: MSB and all authors.

*Critical review of the manuscript for important intellectual content*: All authors.

*Statistical analysis*: MSB, RT, AY.

*Obtained funding*: All authors.

*Administrative, technical, or material support*: All authors.

*Supervision*: MSB, DEG, JDB, JK, TH.

## Acknowledgements

We sincerely thank the families of the brain donors for their invaluable contributions to this research. Without their generosity and willingness to support scientific discovery, this work would not have been possible. We also acknowledge the Lieber Institute for Brain Development Human Brain Repository and collaborating institutions for providing access to these precious samples. Their dedication to advancing our understanding of brain disorders is deeply appreciated.

Additionally, we extend our gratitude to the physicians and staff of the Office of the Medical Examiner, Kalamazoo, Michigan; the University of North Dakota Forensic Pathology Center, Grand Forks, North Dakota; the Office of the Medical Examiner-Coroner, County of Santa Clara, San Jose, California; and the Office of the Chief Medical Examiner for the State of Maryland, Baltimore, Maryland, for their time and efforts to advance tissue donation for neuroscientific research into neuropsychiatric disorders.

