## Supplemental Figures 1-13 for "Convergent molecular signatures across eating disorders and obsessive-compulsive disorder in the human brain"

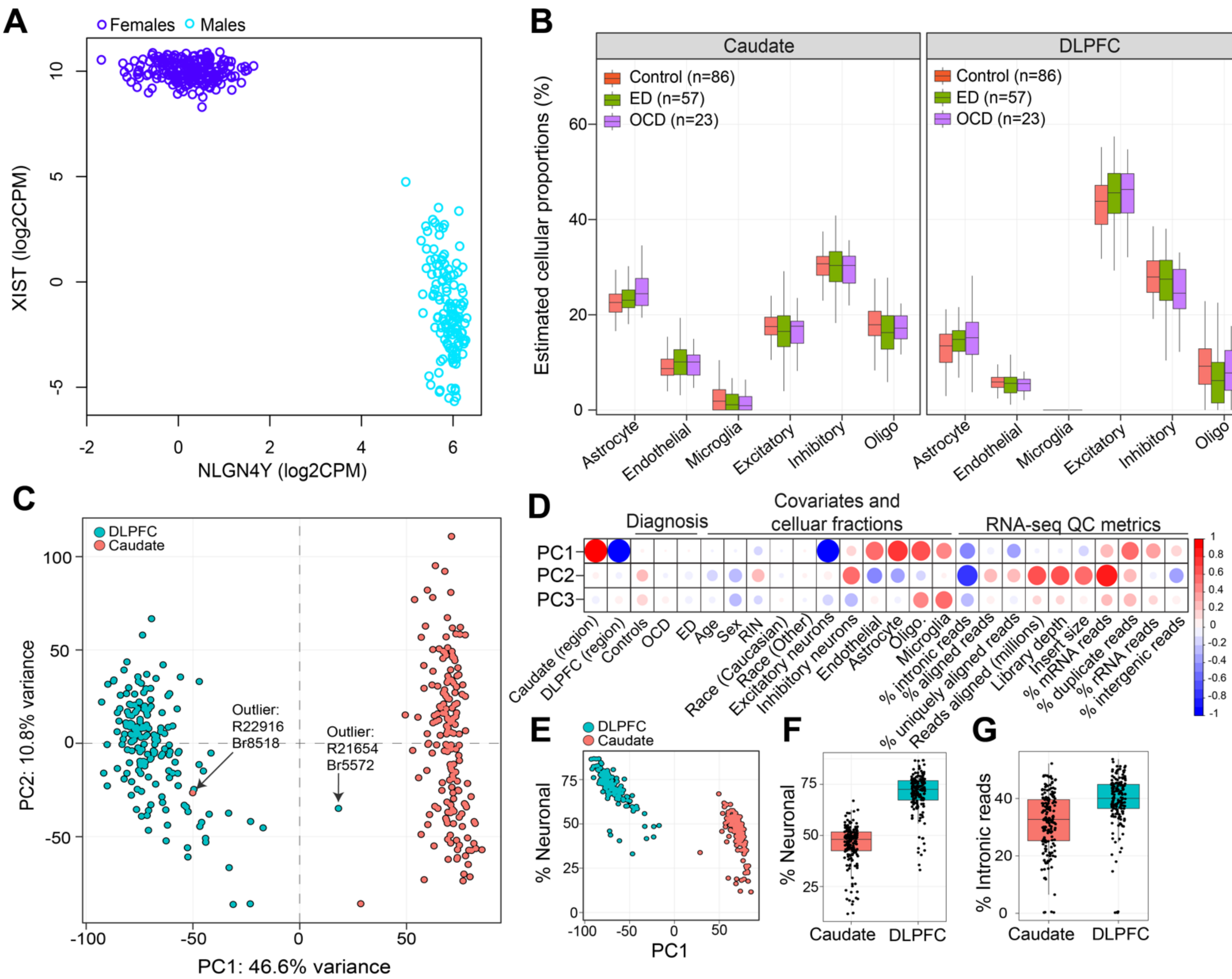


**Supplemental Figure 1: Rigorous quality control measures for RNA-sequencing data.** (**A**) Biological Sex Verification: This panel demonstrates biological sex confirmation through the comparative normalized gene expression of XIST (female-specific, depicted on the y-axis) and NLGN4Y (male-specific, plotted on the x-axis). (**B**) Cellular Composition Deconvolution: Utilizing the MIND R package in conjunction with the Darmanis et al., signature matrix, this panel illustrates the estimated cellular composition. Proportions of each cell type (y-axis) are calculated for all samples across the brain transcriptomes, differentiating between the caudate (left) and the dorsolateral prefrontal cortex (DLPFC) (right). Notably, the caudate is characterized by an increased presence of astrocytes, microglia, and oligodendrocytes, while the DLPFC predominantly consists of neuronal populations (combination of excitatory and inhibitory neurons). (**C**) Principal Component Analysis (PCA): The PCA of VOOM normalized gene expression data showcases the segregation of the DLPFC and caudate transcriptomes. The graph identifies two outlier samples, which have been subsequently excluded from further analysis to ensure data integrity. (**D**) Correlation of Principal Components with External Factors: This plot correlates principal components (y-axis) with a range of clinical, technical, and biological factors (x-axis) determined by Pearson's correlation coefficient, providing insights into the major influences on gene expression variance. (**E**) Neuronal Content Association with PC1: This association graph links PC1 with the percentage of neuronal content in each sample (y-axis; sum of excitatory and inhibitory neurons), highlighting a trend of increasing neuronal proportion. (**F**) Comparative Neuronal Content: A bar graph detailing the relative increase in neuronal content percentage in the DLPFC in comparison to the caudate. (**G**) Intronic Read Proportions: This component delineates the elevated percentages of intronic reads observed in the DLPFC relative to the caudate, which may reflect differences in gene expression regulation or transcript processing between these brain regions.

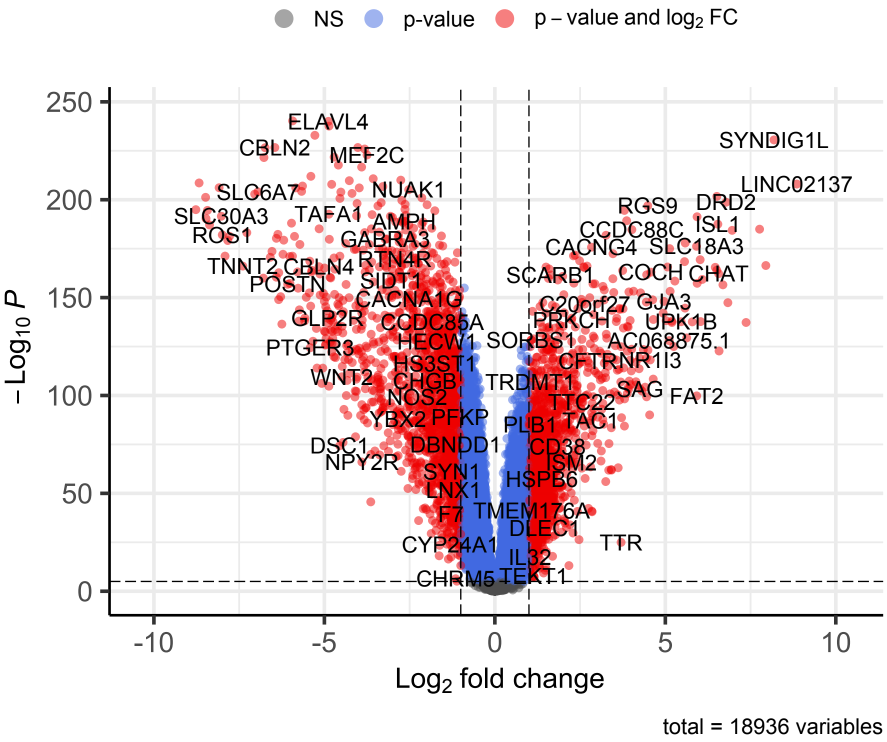


**Supplemental Figure 2. Regional variations in gene expression.** This figure presents a comprehensive differential gene expression analysis between the dorsolateral prefrontal cortex (DLPFC) and caudate. A total of 18,936 genes underwent joint processing and normalization to assess differential expression, incorporating covariates such as neuronal content, biological sex, age, clinical diagnosis, RNA integrity number (RIN), and race. The volcano plot illustrates gene-level log2 fold changes (x-axis) against their corresponding significance levels (-log10 adjusted p-value [FDR <1%], y-axis). We identified 8,653 genes with significantly higher expression levels in the caudate (displayed on the right side of the plot) as opposed to 7,742 genes more significantly expressed in the DLPFC (represented on the left side). Collectively, these findings indicate that approximately ~86% of the genes analyzed manifest differential expression between these two critical brain regions.

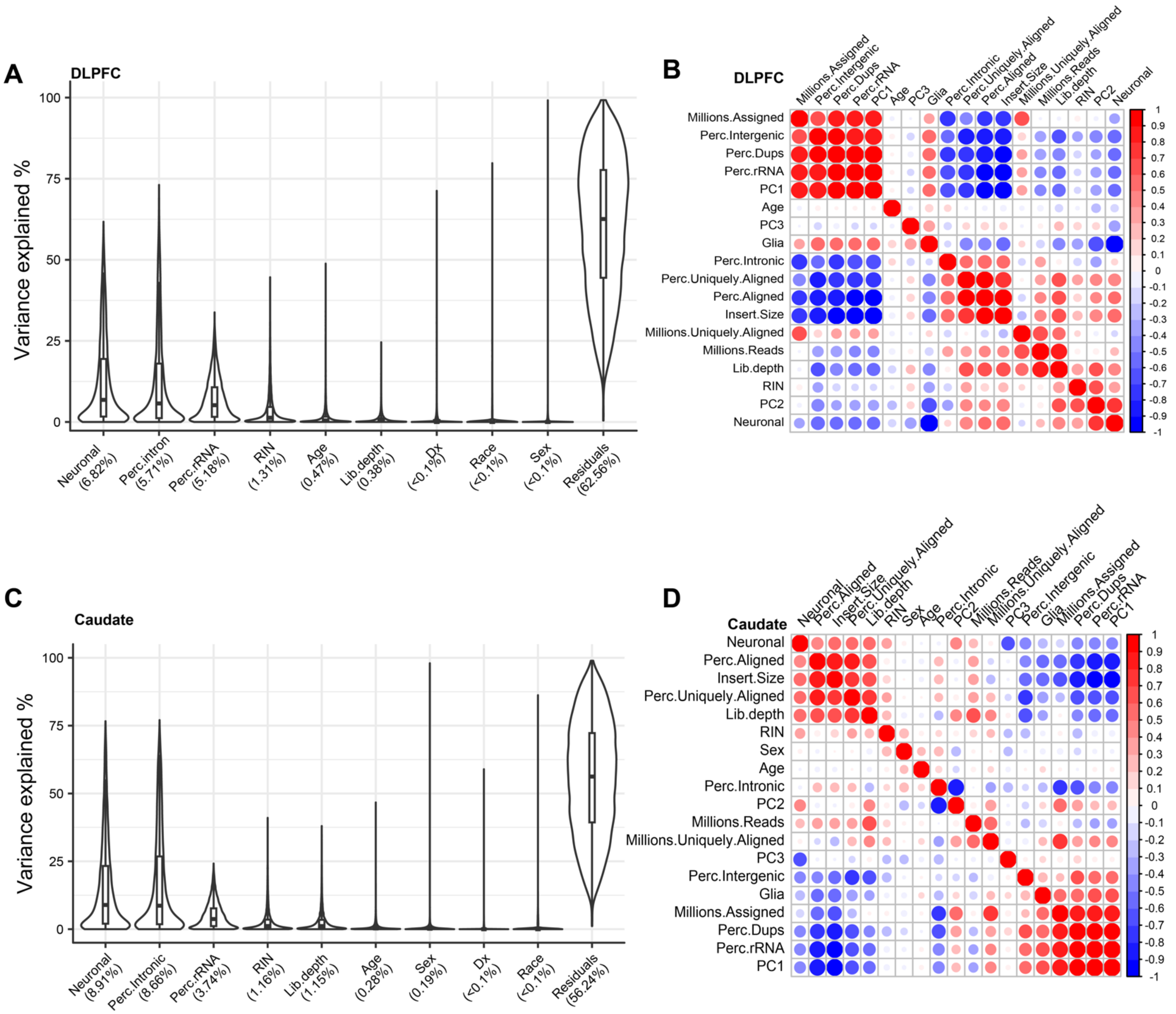


**Supplemental Figure 3. Analysis of variance in gene expression.** (**A**) Variance Decomposition in the DLPFC: This panel employs the variance partition R package to delineate the proportion of gene expression variability in the dorsolateral prefrontal cortex (DLPFC) attributable to an array of biological, technical, and demographic factors. (**B**) Covariate Correlations in the DLPFC: Displayed is a clustered heatmap visualizing pairwise correlations between covariates in the DLPFC, calculated using Pearson's correlation coefficient. This heatmap provides a visual representation of how closely related different variables are in contributing to gene expression variation within the DLPFC. (**C**) Variance Decomposition in the Caudate: Analogous to panel A, this graph quantifies the fraction of RNA-sequencing gene expression variability in the caudate explained by the same set of known variables. (**D**) Covariate Correlations in the Caudate: This is a clustered heatmap akin to panel B, representing pairwise correlations among covariates in the caudate, employing Pearson's correlation coefficient for calculation. The heatmap allows for the assessment of inter-relationships among the covariates with respect to their influence on gene expression in the caudate.

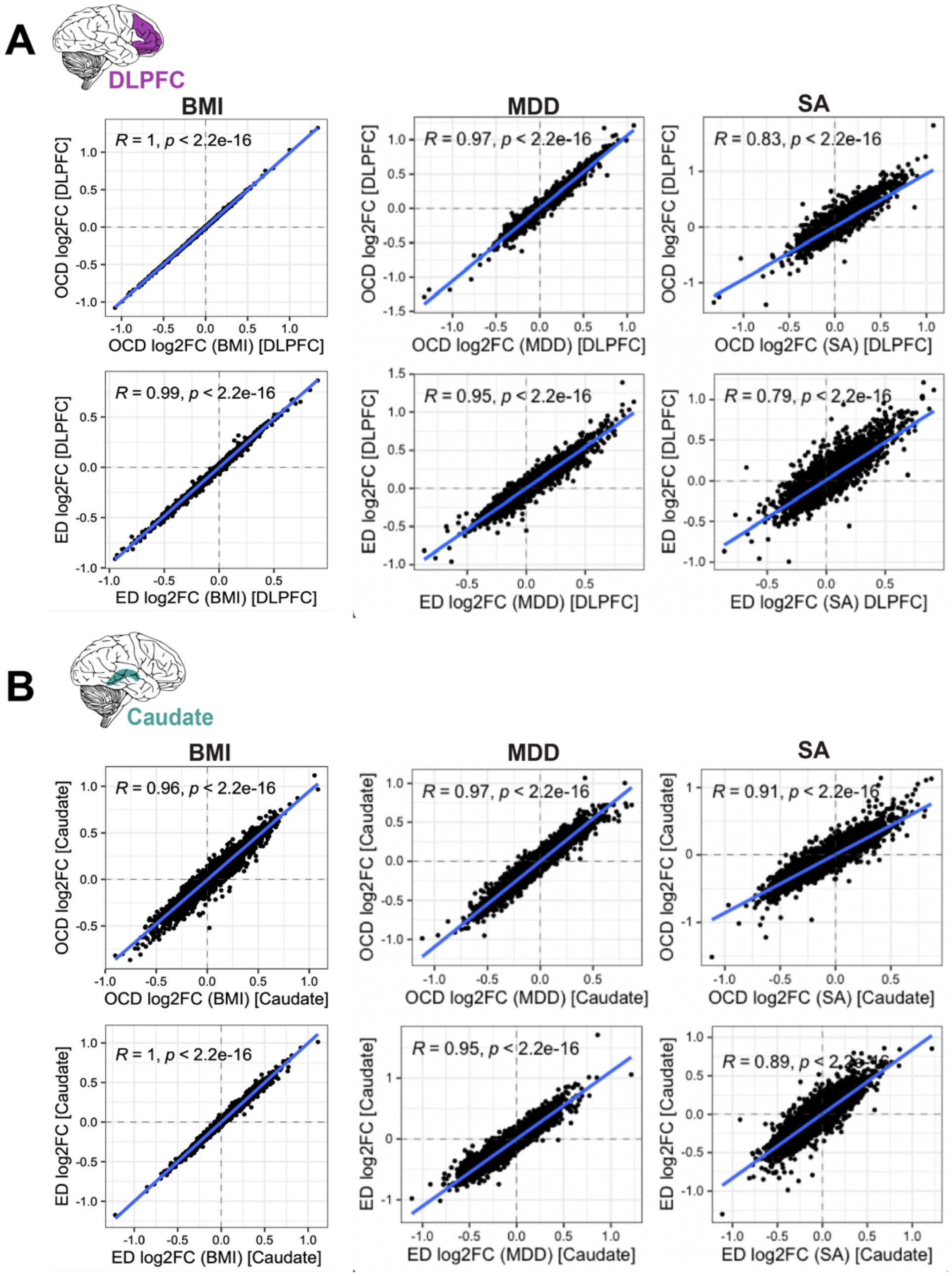


**Supplemental Figure 4. Sensitivity assessment of covariate impact on gene expression.** (**A**) **DLPFC Sensitivity**: This section of the figure examines the sensitivity of differential gene expression in the dorsolateral prefrontal cortex (DLPFC) to body mass index (BMI), comorbid major depressive disorder (MDD) and substance abuse (SA). Substance abuse was categorized as opioid use and/or alcoholism, cannabis use, or polysubstance abuse. We refined our linear model by including these comorbidities as additional variables and contrasted the adjusted log2 fold-changes (FC) against our primary results without these covariates. (**B**) **Caudate Sensitivity**: Following the approach in (A), this segment focuses on the caudate to determine the robustness of gene expression against the added covariates of BMI, comorbid MDD and SA. The comparison of log2 fold-changes (FC) from the adjusted model to the original data set reveals a high degree of correlation, suggesting consistent gene-level effect sizes and direction of effect, confirmed by Spearman's correlation coefficients.

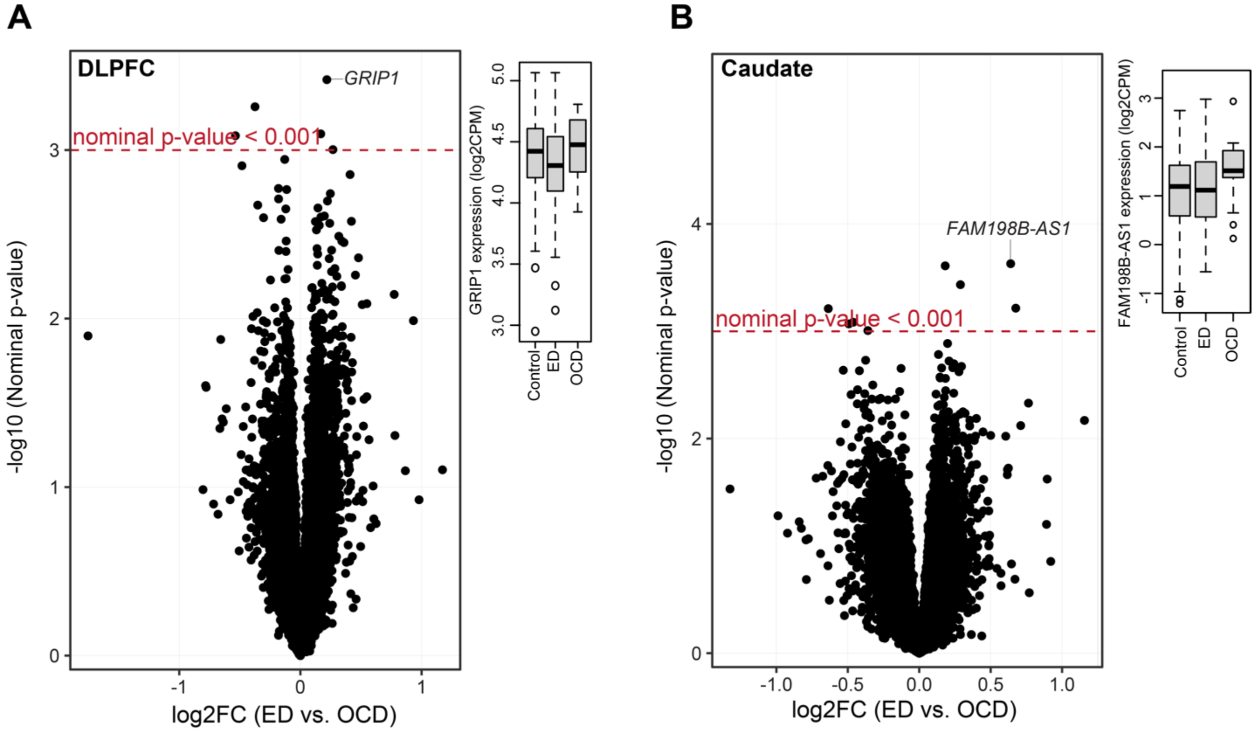


**Supplemental Figure 5. Comparative gene expression analysis between ED and OCD.** (**A**) **DLPFC Expression Differences**: Volcano plots illustrate gene-level significance for the dorsolateral prefrontal cortex (DLPFC), with -log10 nominal p-values on the y-axis plotted against log2 fold-changes (FC) on the x-axis. Despite rigorous analysis, no genes were identified that surpassed the threshold for multiple test correction (FDR < 5%). The plots are marked with a nominal unadjusted p-value threshold line at $p < 0.001$ (dotted line) to emphasize the minimal expression differences between ED and OCD cases. (**B**) **Caudate Expression Differences**: Mirroring the approach in (A), these volcano plots represent gene-level significance in the caudate. As with the DLPFC, no genes met the threshold for multiple test correction. The nominal p-value cut-off is again represented by the dotted line for clarity. To further delineate the negligible effect sizes between ED and OCD cases in both the DLPFC and caudate, boxplots for the most differentially expressed genes are provided. These visualizations highlight that, while some variation exists, the differences do not reach statistical significance.

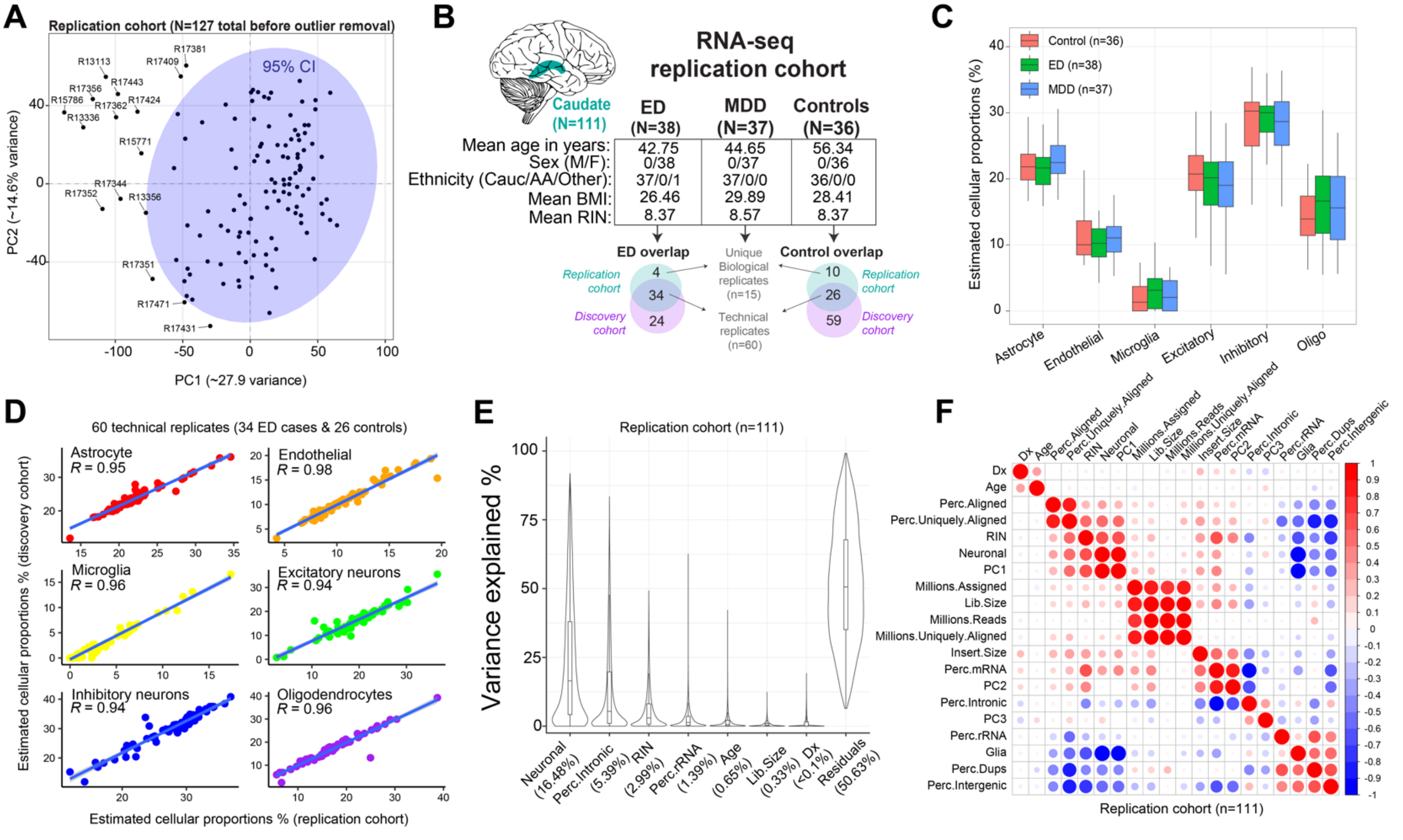


**Supplemental Figure 6. Quality control metrics in the auxiliary replication cohort. (A) Principal component analysis (PCA):** PCA of VOOM normalized gene expression data for 127 samples in the replication cohort. The ellipse indicates a 95% confidence interval (CI) around the grand mean of expression variability, identifying 16 samples that were discarded from subsequent analyses. **(B) Overview of the remaining samples in the replication cohort:** Bulk RNA-seq data encompass 111 donors, examining the caudate nucleus of 38 ED cases, 37 MDD cases, and 36 neurotypical controls. The number of unique biological and technical replicates in common with the discovery cohort are depicted. **(C) Cellular Composition Deconvolution:** Using the MIND R package with the Darmanis et al., signature matrix, this panel illustrates the estimated cellular composition. Proportions of each cell type (y-axis) are calculated for all samples across the caudate transcriptomes. **(D) Replication of estimated cell type proportions between discovery and replication cohorts:** A total of 60 technical replicates were used to show highly similar predicted cell type proportions across repeated measures between the replication cohort (x-axis) and discovery cohort (y-axis). **(E) Variance Decomposition in the Caudate:** This figure quantifies the fraction of RNA-sequencing gene expression variability in the caudate explained by the same set of known variables outlined in the discovery cohort. Note that the replication cohort consists solely of females of similar ethnic backgrounds, and therefore these factors are not included in this analysis. **(F) Covariate Correlations in the Caudate:** Pairwise correlations among covariates in the caudate are calculated using Pearson's correlation coefficient. The heatmap allows assessment of inter-relationships among the covariates and their influence on gene expression in the caudate.

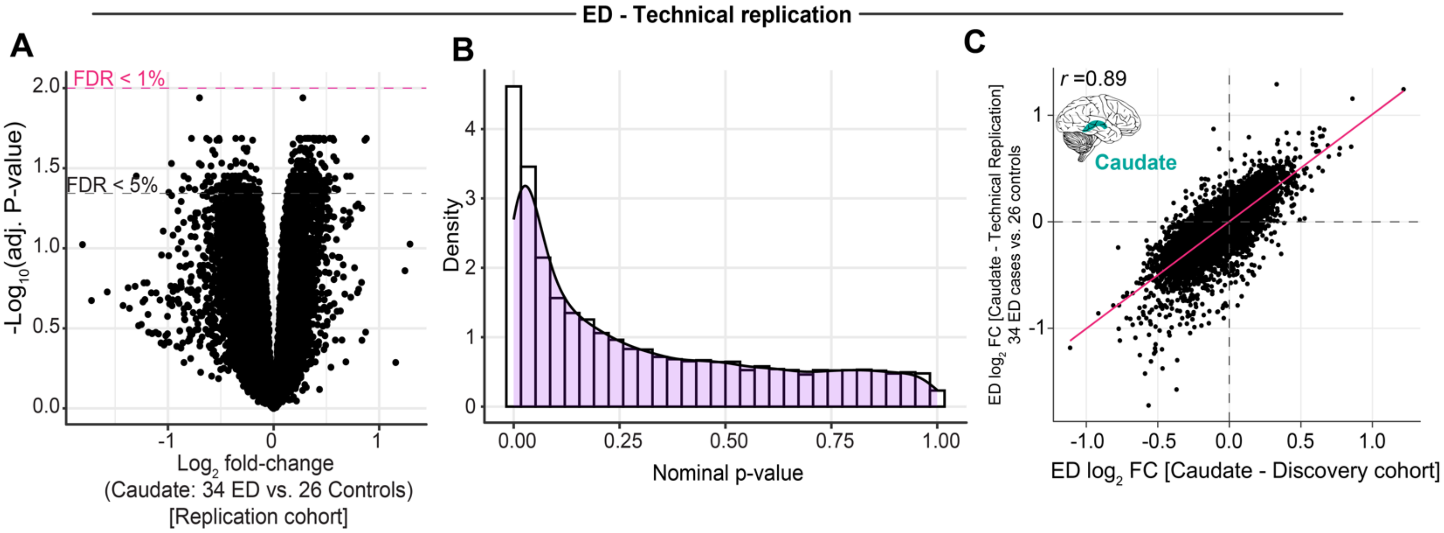


**Supplemental Figure 7. Technical replication of ED transcriptional signatures in the caudate.** (**A**) Differential gene expression analysis in the replication cohort compared 34 ED cases and 26 controls. The volcano plot illustrates gene-level log2 fold changes (x-axis) against their corresponding significance levels (-log10 adjusted p-value [FDR <1%], y-axis). No individual gene met a FDR < 1% threshold, likely due to the limited power of the replication cohort. (**B**) ***P*-value Distribution**: Density plots illustrate an anticonservative *p*-value distributions from differential expression analysis between ED and controls in the caudate. (**C**) **Transcriptome-wide Concordance**: Scatter plots compare log2 fold-change for ED gene-level effects in the discovery (x-axis) and replication cohort (y-axis). Spearman's correlation coefficient (R-values) was calculated to assess concordance. Differential expression accounted for neuronal proportions, RIN, age, sex, and percentage of intronic and rRNA reads. Unlike our analysis in the discovery cohort, we did not include sex or ethnicity as covariates because this replication cohort is composed of only females of European decent.

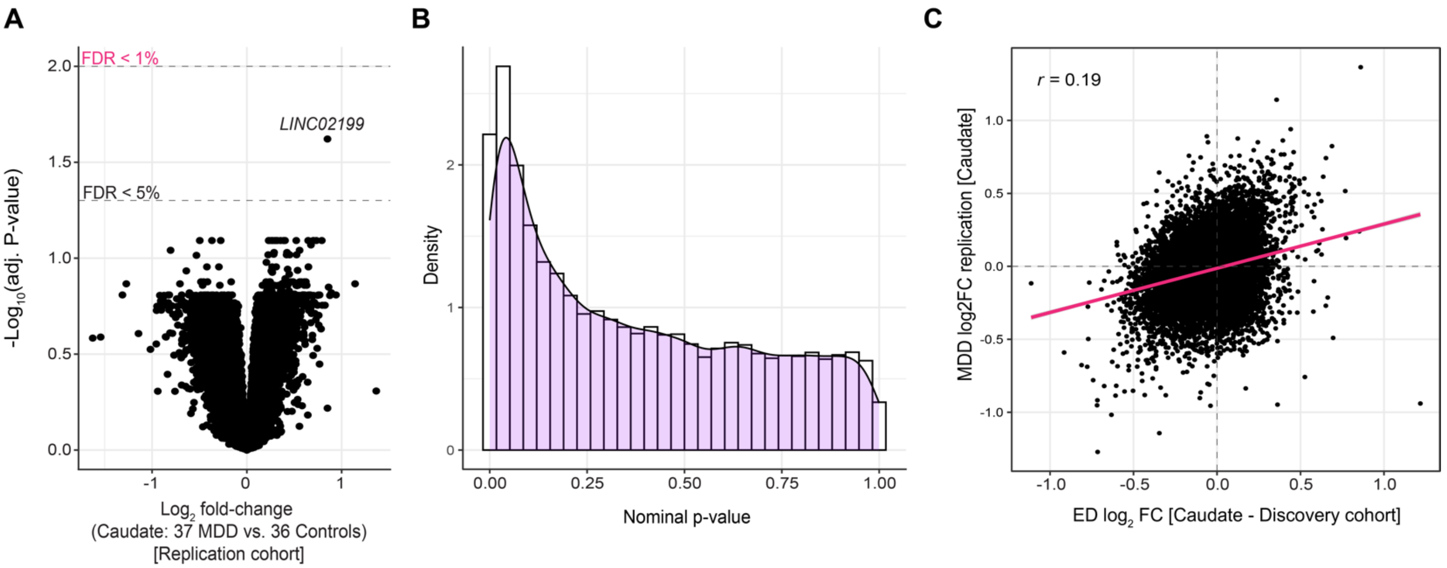


**Supplemental Figure 8. MDD transcriptional signatures in the caudate.** (**A**) Differential gene expression analysis in the replication cohort compared 37 MDD cases and 36 controls (26 technical replicates and 10 unique biological replicates when compared with the discovery cohort). The volcano plot illustrates gene-level log2 fold changes (x-axis) against their corresponding significance levels (-log10 adjusted p-value [FDR <1%], y-axis). No individual gene met a FDR < 1% threshold. (**B**) ***P*-value Distribution**: Density plots illustrate an anticonservative *p*-value distributions from differential expression analysis between MDD and controls in the caudate. (**C**) **Transcriptome-wide Concordance**: Scatter plots compare log2 fold-change between ED gene-level effects in the discovery (x-axis) and MDD gene-level effect sizes in replication cohort (y-axis). Spearman's correlation coefficient (R-values) was calculated to assess concordance.

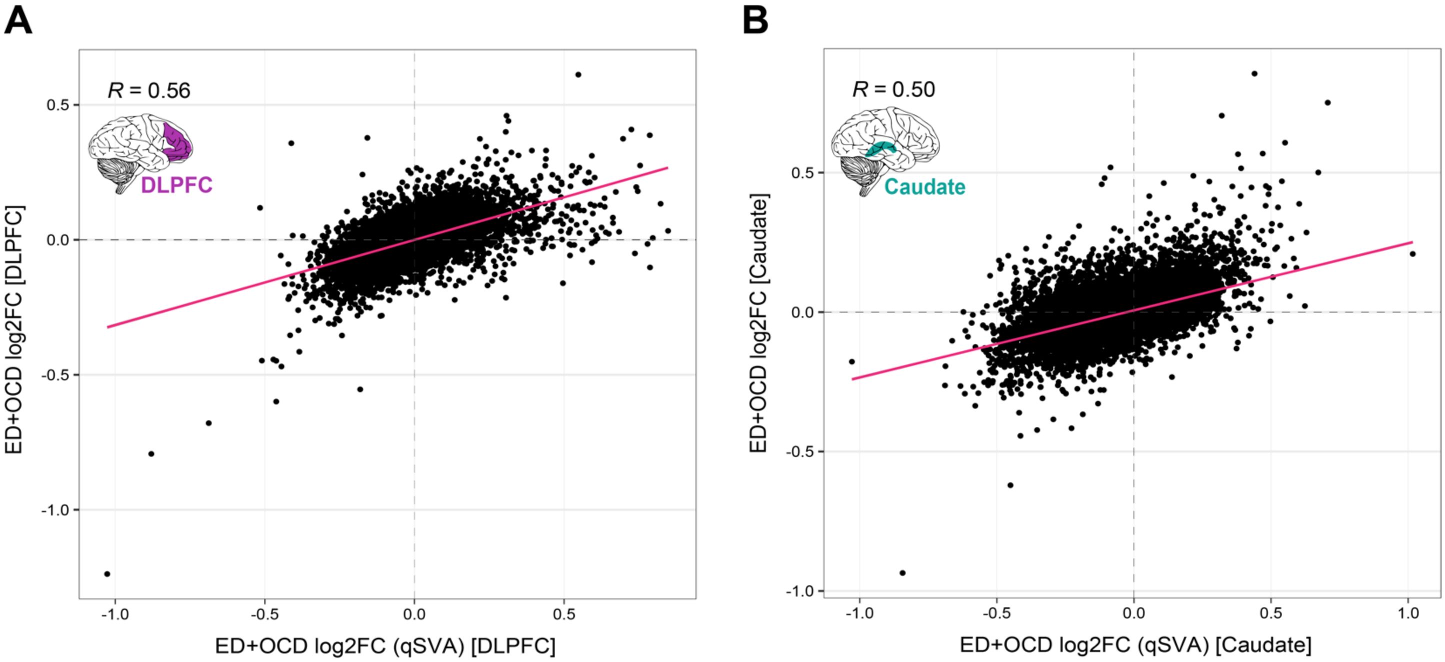


**Supplemental Figure 8. Quality verification using quantitative surrogate variable analysis.** These results assesses the robustness of our differential expression findings through quantitative surrogate variable analysis (qSVA). These analyses aim to adjust for the possible the influence of RNA quality disparities on gene expression. (**A**) **DLPFC RNA Quality Impact Analysis**: A scatter plot displays the correlation between log2 fold-changes derived from our baseline model (y-axis) against those adjusted for RNA quality differences via qSVA (x-axis) in the dorsolateral prefrontal cortex (DLPFC), with a Spearman's correlation coefficient (R) of 0.56. (**B**) **Caudate RNA Quality Impact Analysis**: Similarly, this scatter plot correlates the baseline log2 fold-changes (y-axis) with qSVA-adjusted log2 fold-changes (x-axis) for the caudate, with an R value of 0.50. These sensitivity analyses confirm the dependability of our differential gene expression results, with a moderate to strong correlation between qSVA-adjusted and baseline fold-changes, asserting that RNA quality did not unduly bias our conclusions.

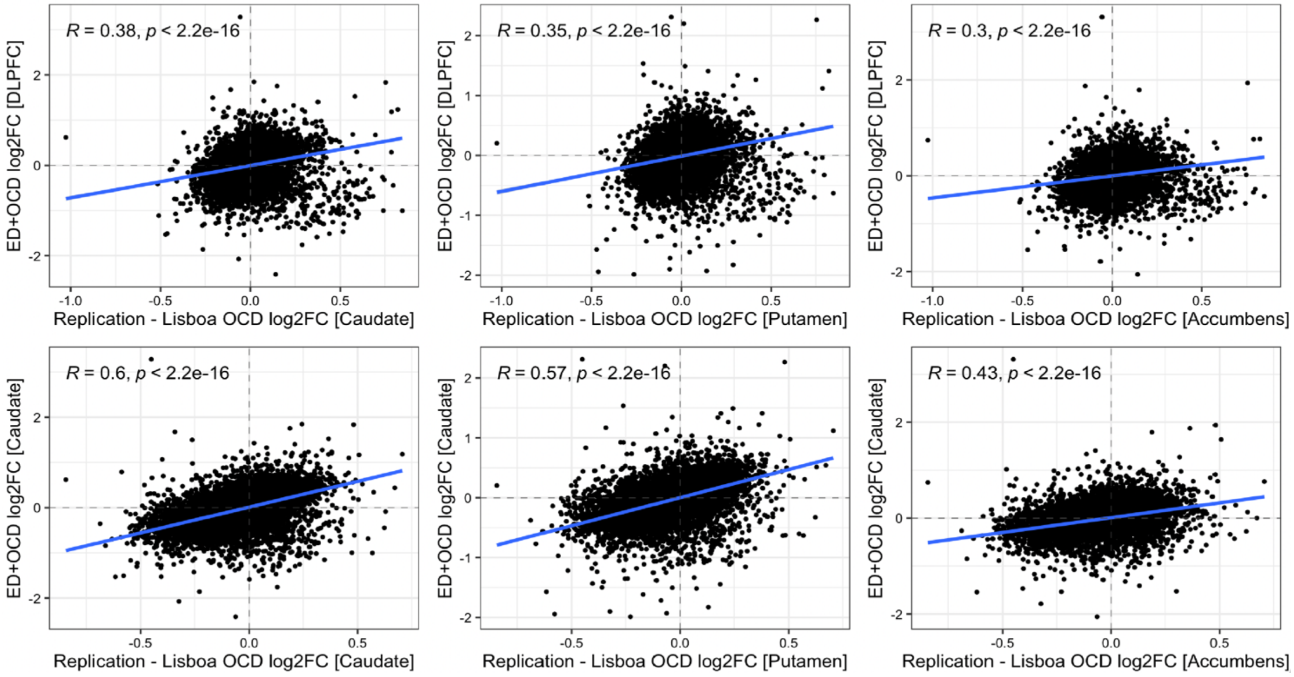


**Supplemental Figure 9: Concordance with an independent OCD brain transcriptome study. Comparative Analysis with Lisboa et al., 2019**: The scatter plots in this figure exhibit the replication efforts of ED+OCD differential gene expression findings with an independent study by Lisboa et al., 2019, which analyzed brain transcriptomes from 7 OCD cases and 8 neurotypical controls. The regions of focus were the caudate, putamen, and accumbens. The datasets from Lisboa et al. were re-analyzed using the same processing methodologies as in our study to ensure comparability. We assessed the concordance of transcriptome-wide log2 fold-change (FC) between our ED+OCD results and those reported by Lisboa et al., employing Spearman's correlation coefficient as a measure of replication strength. The analysis aims to provide an external validation of our findings by demonstrating their reproducibility in a separate cohort and across different brain regions associated with OCD. This cross-validation strengthens the generalizability and reliability of our transcriptomic insights into ED and OCD pathology.

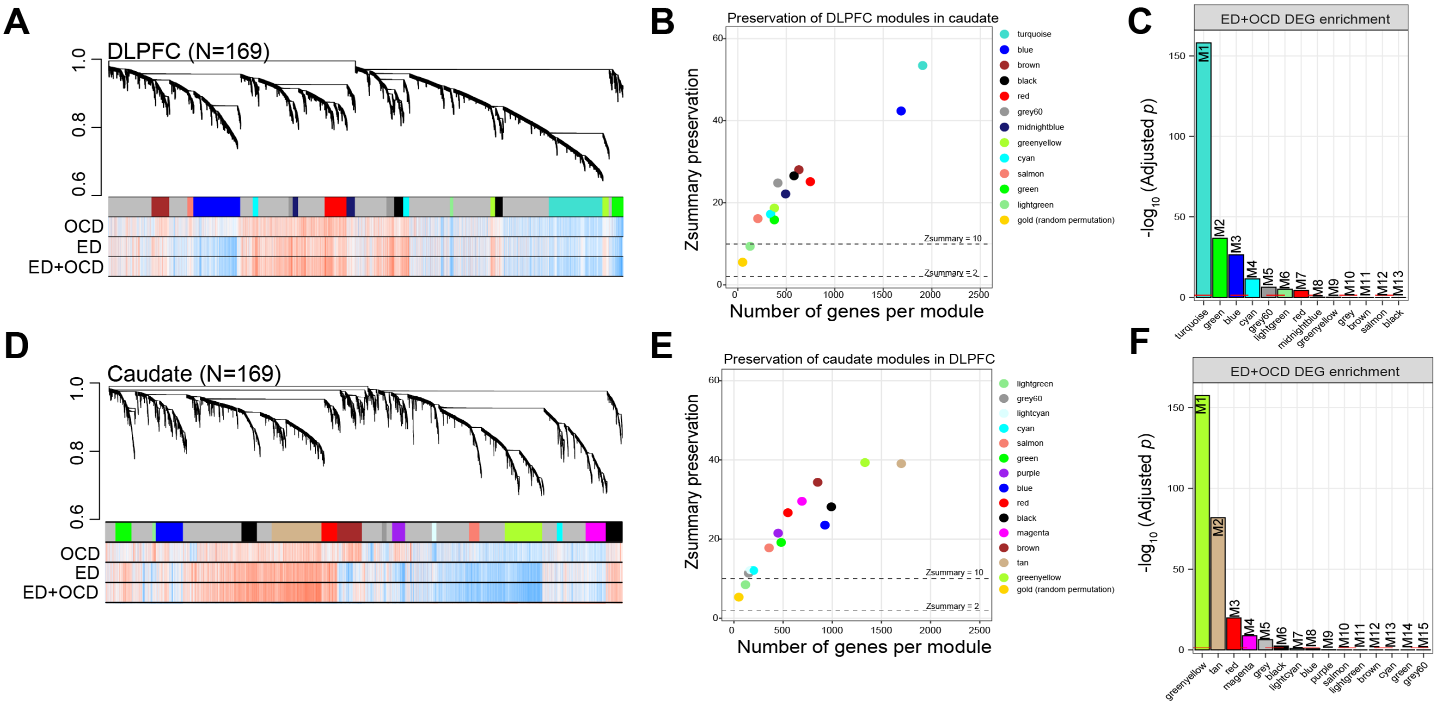

**Supplemental Figure 10. Gene Co-expression Network Analysis and Module Detection.** (**A**) Co-expression Network in the DLPFC: Displays the weighted gene co-expression network analysis (WGCNA) for the dorsolateral prefrontal cortex (DLPFC), identifying 13 distinct modules. The hierarchical clustering dendrogram shows the module structure with genes aggregated based on expression profiles. Adjacent to the dendrogram are color bands representing module membership and gene-trait correlations, where red signifies a strong positive correlation and blue denotes a strong negative correlation with the traits of interest. (**B**) DLPFC Module Preservation in the Caudate: This plot evaluates the preservation of DLPFC-derived gene co-expression modules within the caudate, providing insight into the conservation of gene regulatory networks across different brain regions. A module preservation Zsummary<2 indicates no preservation, 2<Zsummary<10 indicates moderate preservation and Zsummary>10 indicates strong preservation. (**C**) DLPFC Module Enrichment with ED+OCD Differential Expression: The modules originally identified by colors in the DLPFC are relabeled with numbers, with enrichment for eating disorder and obsessive-compulsive disorder (ED+OCD) differentially expressed gene (DEG) signatures being assessed. (**D**) Co-expression Network in the Caudate: Similar to panel (A), this represents the WGCNA for the caudate, elucidating 15 modules. The dendrogram alongside gene-phenotype color bands depicts the co-expression modules and their respective correlations with phenotypic traits. (**E**) Caudate Module Preservation in the DLPFC: This analysis ascertains the retention of gene co-expression modules identified in the caudate when applied to the DLPFC, highlighting the cross-regional module stability. (**F**) Caudate Module Enrichment with ED+OCD DEGs: Modules in the caudate are re-labeled from their initial color identification to numerical labels for clarity, with a focus on the enrichment of ED+OCD DEG patterns within these modules.

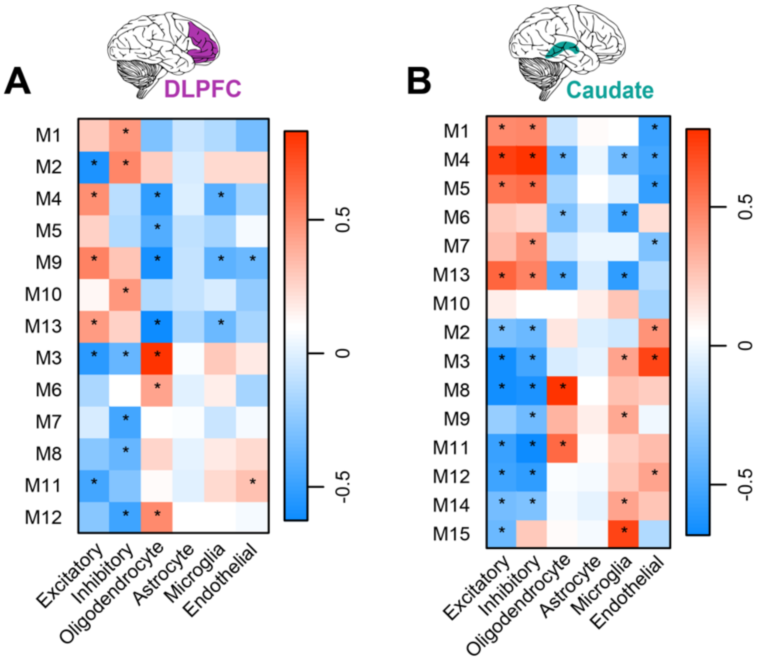


**Supplemental Figure 11. Cell type inferences for co-expression modules.** (**A**) DLPFC Cell Type Correlations: Correlations between the module eigenvalues (y-axis) and the proportions (%) of estimated cell types (x-axis) within the dorsolateral prefrontal cortex (DLPFC). Cell types include astrocytes, endothelial cells, microglia, excitatory and inhibitory neurons, and oligodendrocytes. Associations between module membership and cell type proportions were assessed using Student's asymptotic p-value, with significant correlations denoted by (Bonferroni $p$ <0.05). (**B**) Caudate Cell Type Correlations: A similar approach is applied to the caudate, correlating module eigenvalues (y-axis) with estimated cell type proportions (x-axis). This analysis helps to infer the cellular context of gene expression modules and their potential biological significance. The strength of association and its statistical significance are also tested with Student's asymptotic p-value, indicated by (*P<0.05) where the association is significant.

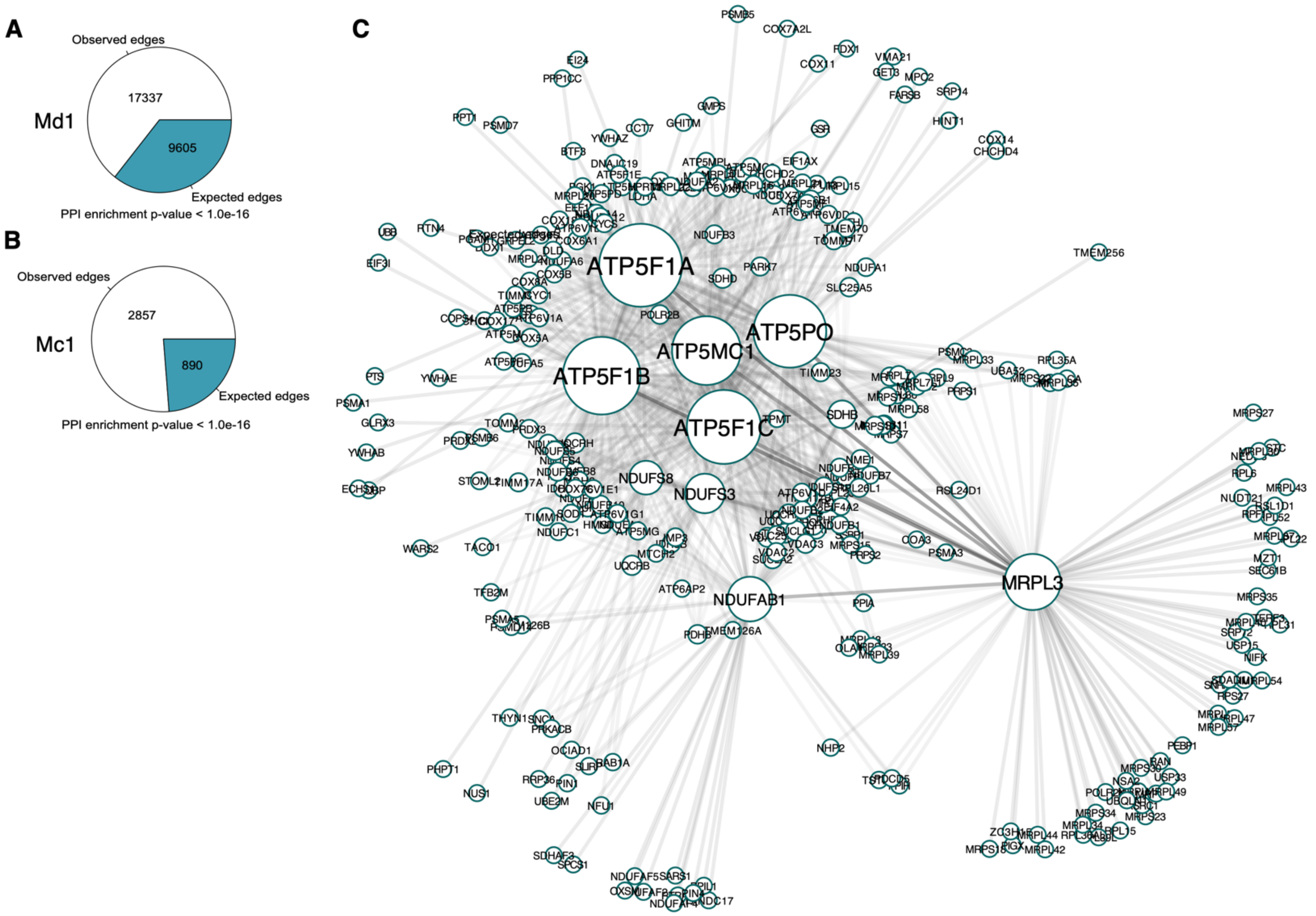


**Supplemental Figure 12. High-confidence protein-protein interaction network.** Expected versus observed edges for protein-protein interaction (PPI) networks for modules (**A**) Md1 and (**B**) Mc1. STRING tests whether the number of observed PPIs are significantly more than expected by chance using a nontrivial random background model. (**C**) This figure features a network visualization of the direct protein-protein interactions among genes from module M1c (associated with the caudate). Generated using the STRING database, the network emphasizes only high-confidence interactions, with a stringent combined string score threshold set at >0.8, ensuring the reliability of the depicted associations. For visualization, the STRING network was imported into Cytoscape.

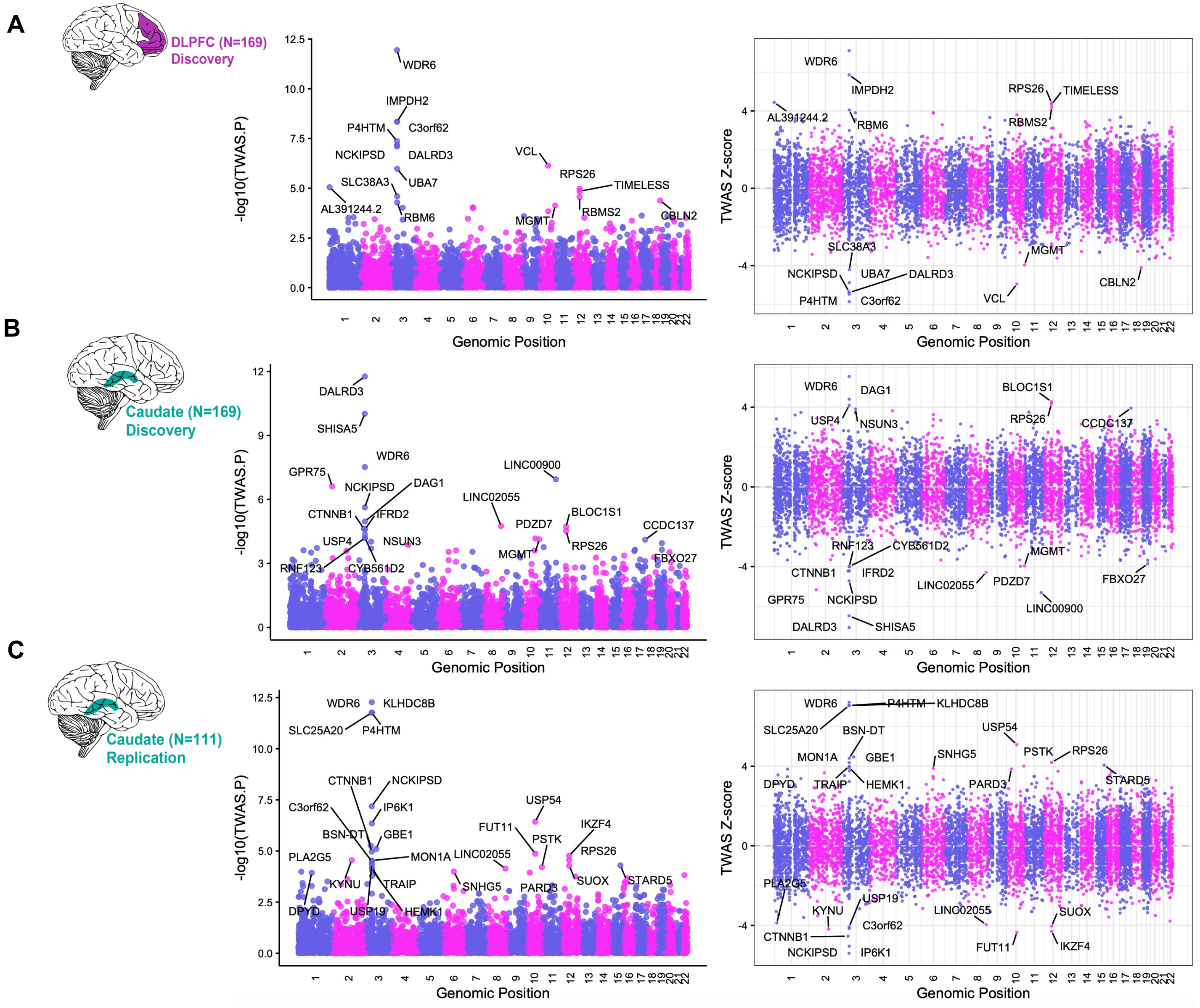


**Supplemental Figure 13**. **FUSION TWAS Results**. This figure presents Manhattan plots summarizing the results of the FUSION Transcriptome-Wide Association Study (TWAS) for ED across three cohorts: (**A**) the Dorsolateral Prefrontal Cortex (DLPFC) discovery cohort, (**B**) the caudate discovery cohort, and (**C**) the caudate replication cohort. Each panel consists of two types of plots – LEFT: A one-sided Manhattan plot on the left, which displays the strength of association between genetically predicted mRNA expression and ED, measured by -log10 TWAS P-values, across the genome. The x-axis represents the chromosomal location of each gene, while the y-axis shows the statistical significance of the association. RIGHT - A two-sided Manhattan plot on the right, which visualizes TWAS Z-scores to indicate both the direction and strength of the associations. Positive Z-scores reflect upregulation associated with increased ED risk, while negative Z-scores indicate downregulation linked to increased risk. All labeled points in both plots represent genes whose associations pass a significance threshold of FDR < 5%, highlighting those that are most likely to have a true effect on ED risk.
